# Outpatient clonal propagation propelled rapid regional establishment of an emergent carbapenem-resistant *Acinetobacter baumannii* lineage ST499^Pas^

**DOI:** 10.1101/2022.06.21.22276711

**Authors:** Juan J. Calix, Maria C. Stockler de Almeida, Robert F. Potter, Meghan A. Wallace, Carey-Ann D. Burnham, Gautam Dantas

**Affiliations:** Division of Infectious Diseases, Washington University in St. Louis School of Medicine, St. Louis, Missouri 63130, United States of America; The Edison Family Center for Genome Sciences and Systems Biology, Washington University School of Medicine, St. Louis, Missouri 63130, United States of America; Department of Medicine, University of Sao Paolo, Sao Paolo, Brazil; Department of Pathology and Immunology, Washington University School of Medicine, St. Louis, Missouri 63130, United States of America; Department of Molecular Microbiology, Washington University School of Medicine, St. Louis, Missouri 63130, United States of America; Department of Biomedical Engineering, Washington University in St. Louis, St. Louis, Missouri 63130, United States of America

**Keywords:** *Acinetobacter baumannii*, molecular epidemiology, healthcare-associated infections, drug resistance

## Abstract

Combating the evolving health threat posed by carbapenem-resistant *Acinetobacter baumannii* (CR*Ab*) requires knowing how this non-commensal organism establishes regional pools and propagates among at-risk hosts. We report a 2017-2019 surge of CR*Ab* among patients receiving care in a USA multicenter system. This surge occurred during a period of sustained reduction in hospital-acquired CR*Ab* infections and coincided with marked reduction of CR*Ab* cases associated with distinctly more resistant antibiotypes. Isolate whole genome sequencing revealed surge isolates belonged to an emergent Pasteur scheme sequence type 499 (ST499^Pas^). Detailed query of health records guided by isolate genome comparative analyses revealed multiple clonal clusters linked to various outpatient healthcare settings (i.e., long term healthcare facilities, surgical and wound clinics, and other unidentified factors) but no evidence of a shared intrahospital source. We show that emergent CR*Ab* lineages can rapidly establish a regional presence even without gains in breadth of antibiotic resistance and negligible contribution from sustained intrahospital transmission. The emergence of ST499^Pas^ despite regional eradication of other CR*Ab* lineages shows how control efforts could be sidestepped via outpatient epidemiological niches. We also establish an approach to investigate the propagation of CR*Ab* lineages that can inform subsequent local surveillance efforts outside of hospital settings.

## Background

Recognition of carbapenem-resistant *Acinetobacter baumannii* (CR*Ab*) as a top priority in the battle against multidrug resistant infections[1] has prompted efforts to interrupt the propagation of this incidental pathogen among at-risk human hosts. CR*Ab* infections disproportionately impact critically-ill individuals, so surveillance efforts have generally focused on intrahospital microbial reservoirs and outbreaks of hospital-acquired (HA) infections[2, 3]. However, reports of CR*Ab* acquisition occurring outside of hospital environments[4–6], raise concerns that outpatient reservoirs could sustain the CR*Ab* epidemic, even if intrahospital transmission is eradicated.

Long-term care facilities and acute care hospitals (herein collectively referred as “LTFs”) are implicated as nonhospital “hubs” communicating multidrug resistant organism (MDRO) outbreaks between hospitals[6–11]. Evolutionary adaptations to colonize human gut, skin, genitourinary or upper respiratory niches may facilitate the flow of MDRO pathobionts (e.g., Enterobacterales, *Staphylococcus, Enterococcus* etc.) among these non-hospital environments. Clinically-relevant lineages of environmental MDROs like CR*Ab,* however, are not routinely identified in these commensal niches[3, 12–14] and their propagation is thought to more heavily rely on microbial reservoirs on nosocomial surfaces[3]. Given a presumptive lack of mobility, it remains controversial whether LTFs and other nonhospital reservoirs can spur sustained occurrence of CR*Ab* disease, independent of stable intrahospital pools.

*Ab* is a genetically diverse species capable of causing many types of infections[3, 15, 16], but only a few global *Ab* lineages like the Pasteur scheme multilocus sequence types 2 and 79 (ST2^Pas^ and ST79^Pas^) have historically accounted for most healthcare-associated CR*Ab* infections[17]. Acquisition of antimicrobial resistance and the ability to colonize nosocomial surfaces likely contribute to these lineages’ fitness in healthcare environments[3]. However, *Ab* demonstrates a high degree of genomic plasticity and access to a deep gene pool[18], which could conceivably foster other lineages incidentally equipped to exploit changing trends in medical practices.

Here, we investigated CR*Ab* propagation behaviors via detailed characterization of cases in a large USA healthcare system experiencing a sustained decrease of HA cases (i.e., cases with index cultures identified >48 hours after hospital admission) but a steady occurrence non-HA (nHA) cases between 2012-2019[16]. The lack of in-hospital outbreaks since 2012 affords an opportunity to study CR*Ab* propagation with limited contribution from intrahospital pools, so we employed whole genome sequencing (WGS) of clinical isolates to describe the regional nonhospital arena contributing to CR*Ab* cases. We also tested the hypothesis that CR*Ab* lineages responsible for HA cases pre-2012 are also driving the recent persistence of nHA cases.

## Methods

### Study location and period

This study was approved by the Washington University Institutional Review Board (IRB# 201707046 and 201707049) and was performed in five hospitals (herein denoted as BJC1-5) in the affiliated BJC HealthCare System (BJC) from January 1, 2007 to December 31, 2019. BJC is a large integrated inpatient and outpatient healthcare system serving St. Louis, Missouri, USA and surrounding areas. The BJC Epic electronic medical record (EMR) system integrates records from 27 of the 29 hospitals in the area, which allowed for review of an individual’s microbiological results, hospitalization notes and outpatient appointments in both BJC and non-BJC systems. Study BJC hospitals and affiliated clinics use a central BJC clinical microbiology laboratory (BJC-CML).

### Retrospective case identification and definitions

The BJC Clinical Data Repository (CDR) was used to identify cases associated with isolates identified as *Acinetobacter* according to automated biochemical methods or matrix-assisted laser desorption/ionization and time of flight mass spectroscopy (MALDI-TOF MS). To account for the unreliability of biochemical methods in distinguishing species within the *Acinetobacter calcoaceticus-baumannii complex* (Acbc), isolates reported as “*Acinetobacter baumannii*” or “*Acinetobacter calcoaceticus-baumannii* complex” were binned as “*Acbc* cases”. However, because carbapenem resistance (CR) is rarely reported in non-*baumannii* species and all genotyped carbapenem-resistant isolates in this and a previous U.S. study[19] were confirmed as *Ab,* all CR isolates were labeled CR*Ab.* Case clinical data was obtained from the BJC CDR and by review of EMR, and cases lacking carbapenem susceptibility data (n=98) and hospital outbreak surveillance cultures (n=54) were excluded. Remaining cases were defined by the first culture containing an *Acinetobacter* isolate per patient (“index culture”) and classified into five categories according to tissue source: “respiratory”, “skin and soft tissue/musculoskeletal” (SST/MSK), “urinary”, “blood” (isolates obtained from blood, central lines, or other endovascular devices or grafts), or “other.” Nearly all 2017-2019 CR*Ab* cases met “healthcare-associated” criteria[20]. So to better resolve changes in epidemiology, we defined cases as “hospital-acquired” (HA) if index culture was collected ≥48 hours after hospital admission and prior to discharge, or “nonhospital-acquired” (nHA) for all others, as done before[16].

### Clinical isolate banking and definitions

We performed a prospective, convenience banking of *Acinetobacter* isolates identified in the BJC-CML between July 1, 2017, and May 31, 2019 (**Figure S1A**). Isolates identified as an *Acinetobacter* species according to MALDI-TOF MS were eligible for inclusion. If more than one morphologically distinct colony on culture plate was identified as *Acinetobacter,* both colonies were stored. At the end of the banking period, isolates were matched to their respective clinical metadata and processed for genomic analysis. The earliest isolate belonging to a Pasteur scheme multilocus sequence typing (MLST) sequence type (ST) per person was denoted the “index isolate”, with subsequent isolates denoted as “non-index isolates.” Multiple index isolates with unrelated STs could be obtained from a single individual. Herein, banked isolates are named by number (e.g., “isolate 212” denotes WU_MDCI_Ab212, etc.).

### Antibiotic susceptibility testing (AST) and antibiotyping

AST was performed in a CLIA and CAP accredited clinical microbiology laboratory, and interpreted per Clinical and Laboratory Standards Institute guidelines[21]. Per BJC-CML protocols, all *Acinetobacter* isolates are routinely tested by Kirby-Bauer disk diffusion on Mueller-Hinton Agar, for susceptibility to ceftriaxone (CRO), ceftazidime (CAZ), cefepime (FEP), meropenem (MEM), piperacillin-tazobactam (TZP), ampicillin-sulbactam (SAM), trimethoprim-sulfamethaxazole (SXT), gentamicin (GM), and ciprofloxacin (CIP). MEM non-susceptible isolates are reflex tested with tobramycin (TOB), amikacin (AMK), imipenem (IPM), doxycycline (DOX) and minocycline (MIN). Zone of clearance (ZOC) results were obtained from the BJC CDR, and index isolates missing AST results were tested when recovered from frozen cultures. Isolates non-susceptible to either IPM or MEM were defined as “CR”. All cases and isolates were assigned an antibiotype, i.e., ast-1 thru ast-5, according to their AST results and the algorithm shown in **Figure S1B.** Non-banked cases missing any AST result in the algorithm, were labeled “non-typeable” (nt).

### Whole-genome sequencing (WGS) and comparative analysis

A full description of the well-established processing pipelines[22] used for genomic analyses (**Figure S1)** is provided in the **Supplementary Text**. All sequence files are available under NCBI BioProject PRJNA739144 (BioSample accession numbers SAMN19774044-4250) (**Data S2**). Genome sequences of *Ab* isolates from other geographical regions were obtained from NCBI for global comparative analyses (**Data S3**). In a multi-step clonality analysis, “presumptive” clonal clusters were identified using pairwise core genome single nucleotide polymorphisms (cgSNP) and whole genome average nucleotide identity (wgANI). Clustered isolates that shared a common ancestor per subsequent intra-ST phylogenetic analysis were binned into “confirmed” clonal clusters labeled with their respective ST and a number (i.e., “cluster 499-1”). Patients in each cluster were labeled according to when an isolate displaying the pertinent antibiotype was first identified in their clinical timeline (see below), where patient A being the earliest.

### Visualizing clinical timelines and suspected transmission sites

Clinical care received by patients in clusters was detailed by extensive review of EMR. Dates for seven “encounter” types as defined in **Table S1,** were logged. The “wound care/surgical clinic” encounter type was chosen because of the predominance of SST/MSK specimens in a recent BJC study[16]. We defined facilities as “suspected transmission sites” if either (a) a CR*Ab-*positive patient shared an encounter wither a different patient within the same cluster with a subsequently positive culture; or (b) a shared encounter preceded positive culture for at least two patients in a cluster. Other shared features incidentally identified in our analysis, are noted in Results.

### Statistical analysis

Univariate analyses were performed with SPSS v25 (IBM, USA) or R software v 3.6.2[23]. Chi-squared or independent t-test was performed for comparing categorical or continuous variables, respectively. Statistical significance was defined as *p* values <0·05.

## Results

### Shifts in CR*Ab* epidemiology in BJC

Of 2,157 *Acbc* cases identified in five BJC hospitals in 2007-2019, 2,059 index cultures with carbapenem susceptibility results were eligible for analysis, including 928 (45·1%) CR*Ab* cultures (**Data S1**). The annual percent of CR*Ab* cases being nHA increased from 30-45% in 2007-2009 to 60-75% in 2014-2019 **(Figure 1A**). Additionally, latter CR*Ab* isolates were more susceptible to non-carbapenem antibiotics (**Table S2**), with the prevalence of the most narrowly resistant antibiotype, ast-5, increasing from 1·8% of CR*Ab* cases (13/726) prior to 2017 to 73·6% (23/40) in 2019 (**Figure 1B**). This coincided with the near disappearance of more broadly resistant antibiotypes that prevailed pre-2012, when CR*Ab* cases were predominantly HA (**Figure 1**).

**Figure 1.**
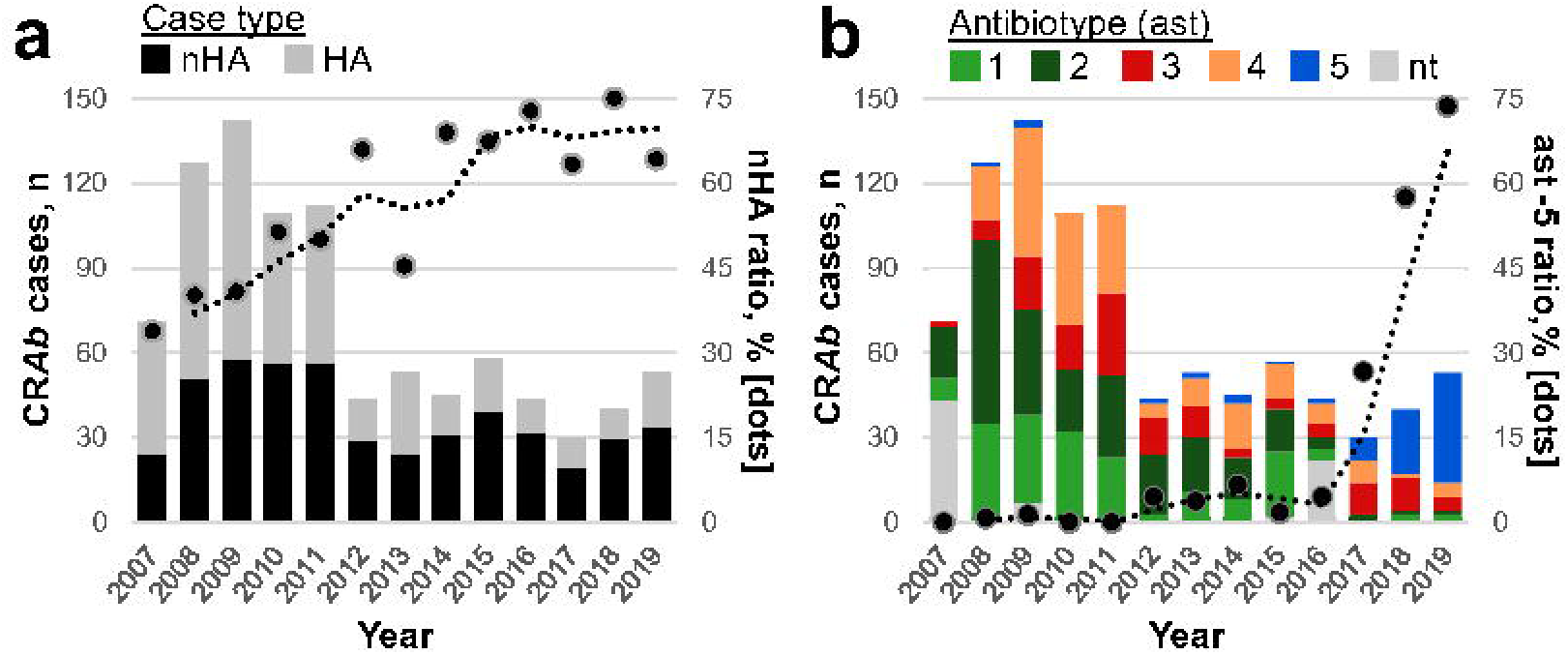
Recent BJC carbapenem-resistant *Acinetobacter* (CR*Ab*) cases are principally non-hospital acquired (nHA) and associated with isolates displaying a unique antibiotype (ast). **(a)** Annual CR*Ab* cases that were hospital-acquired (HA) and nHA (gray and black bars, respectively) and annual percentage of cases that were nHA (“nHA ratio”, dots). **(b)** Annual CR*Ab* cases associated with index isolates exhibiting each antibiotype (colored bars) and annual percentage of cases associated with ast-5 isolates (“ast-5 ratio”, dots). Y-axes values in all panels represent rolling two-year average for respective ratios. nt, non-typeable due to missing AST data (see Figure S1B). Raw values are listed in Data S1.

### ST499^Pas^ CR*Ab* lineage displaying a distinguishable antibiotype predominates BJC

To better understand this shift in CR*Ab* epidemiology, we performed WGS on 207 isolates representing 53·5% (177/331) and 62·0% (54/87) of total *Acinetobacter* and CR*Ab* cases, respectively, identified in BJC between July 2017 and May 2019. Except for a higher likelihood of being banked in earlier calendar quarters, characteristics of banked and non-banked cases were alike (**Table S3 and S4**). wgANI confirmed 90 index and 20 non-index *Ab* isolates from 87 cases (**Data S2**), including cases where strains from distinct Pasteur scheme multilocus sequence types (ST^Pas^) were co-isolated on the same (isolates 32/33 and 204/205) or subsequent dates (isolates 215/223). All genotyped CR isolates belonged *Ab* STs: ST499^Pas^, ST406^Pas^, ST2^Pas^, ST79^Pas^, ST78^Pas^ and ST1^Pas^ (n= 35 [38·9% of total Ab index isolates], 13 [14·4%], 5, 3, 2, and 1 respectively) **(Figure 2A**). Isolates within these STs largely shared lineage-associated beta-lactamase gene repertoires, according to antimicrobial resistance gene analysis (**Figure S2A**). Of 35 isolates exhibiting the ast-5 antibiotype, 31 (88·6%) were ST499^Pas^. The other four (i.e., isolates 39, 165, 233, and 249) were outlier ST406^Pas^ isolates lacking *ant(2”)-Ia*/aadA2 and *sul1,* the genes putatively conferring aminoglycoside and sulfonamide resistance, respectively, in all other ST406^Pas^ genomes and the outlying ST499^Pas^ isolate 212 (**Figure 2** and **Data S4**).

**Figure 2.**
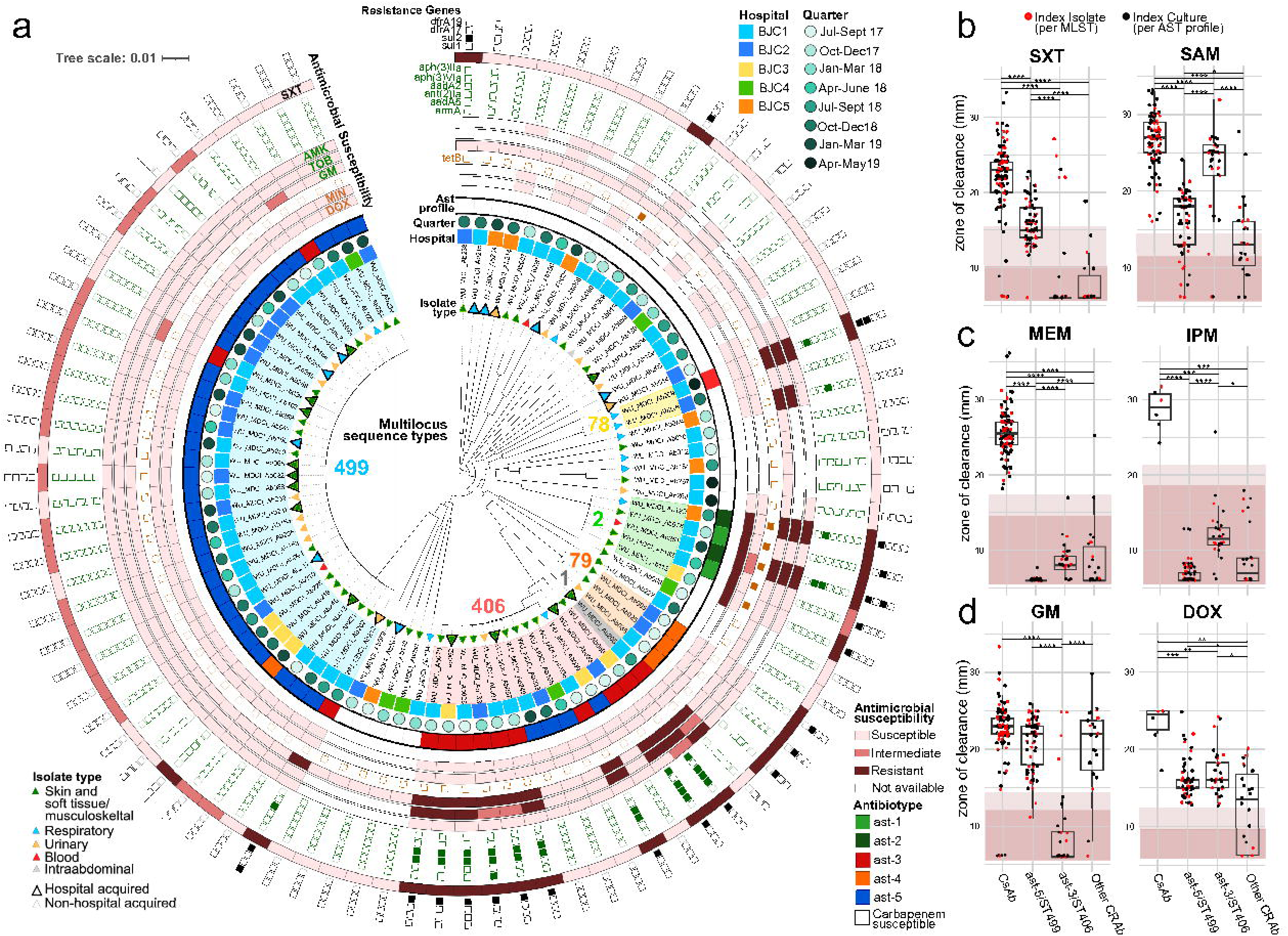
Unique ast-5 antibiotype is linked to prevalent CR*Ab* lineage BJC ST499^Pas^. **(a)** Maximum likelihood phylogenetic tree of BJC *A. baumannii* index isolates according to core genome alignment. ST^Pas^ containing CR*Ab* isolates are highlighted and labeled. Inner rings represent isolate metadata according to the corresponding keys surrounding the tree. Outer rings show AST results grouped by class and flanked by boxes denoting the presence (filled box) or absence of listed genes putatively conferring class resistance. For clarity, only genes exclusive to resistant isolates are included. Isolate metadata are listed in **Data S2**, and all identified resistance genes are listed in **Data S4**. **(b-d)** Antibiotic susceptibility according to zone of clearance, of genotyped *Ab* index isolates (red dots) and non-genotyped *Acbc* index cultures (black dots) identified between January 2017 and December 2019. Non-genotyped CR*Ab* were grouped with ST499^Pas^, ST406^Pas^ or “other” isolates according to antibiotype. Backgrounds highlight ranges for “resistant” (dark) and “intermediate” (light) susceptibility, per CLSI guideline interpretation. Box-plot center lines denote medians, while box limits denote upper and lower quartile values (listed in **Table S5).** Whiskers denote 1.5x interquartile range. Medians compared according to Mann-Whitney test with Bonferroni adjustment for multiple comparisons. *, *p*<0·05; **, *p*<0·005; ***, *p*<0·0005; ****, *p*<0·00005.

Comparison of the AST ZOC data of 2017-2019 *Ab* cases (**Figure 2B-2D, Figure S2B, Table S5**) revealed that genotyped ST499^Pas^ and non-genotyped ast-5 isolates composed a distinctively homogenous group. As a group they displayed (a) intermediate SXT susceptibility lower than most Cs*Ab* isolates and ST406^Pas^ outliers that lacked *sul1* (resulting in the variation reported in their SXT AST results, **Figure 2A and 2B**); (b) ZOC to tetracyclines and aminoglycosides comparable to other susceptible CR*Ab*; and (c) characteristic ZOC distribution to each β-lactam antibiotic, including high resistance to MEM/IPM (**Figure 2C**) but intermediate susceptibility to CAZ/FEP **(Figure S2B**). Altogether, this supported that ST499^Pas^ is responsible for the recent surge of ast-5 isolates observed in BJC (**Figure 1B**).

### ST499^Pas^ clonal clusters were linked to regional nonhospital settings

A global phylogenetic analysis incorporating 510 genomes of *Ab* isolates from other regions revealed that BJC ST2^Pas^ and ST406^Pas^ isolates represented multiple subgroups within their respective global clades (**Figure S3**), consistent with sporadic introductions of these ST into our cohort. In contrast, a single phylogenetic cluster contained all BJC ST499^Pas^ isolates except for the outlying isolate 212, which was obtained from an individual transferred from an unaffiliated hospital in the Southeastern USA. ST499^Pas^ isolates were regularly isolated throughout 2017-2019 from various tissues sources and in four of the five study hospitals, and only 13 of 35 cases met HA criteria (**Figure 2**). So, despite sharing a recent common ancestor, ST499^Pas^ cases unlikely resulted from a spatiotemporally hyperlocalized outbreak.

To understand how BJC ST499^Pas^ emerged in our region, we assigned presumptive clonal relationships between isolates according to cutoffs of cgSNP distance ≤5 and wgANI ≥99·997% (**Figure 3A-B**). The use of more inclusive cutoffs (i.e., cgSNP distance 10-30 and wgANI 99·990-99·995%) yielded minimal changes to presumptive clusters in sensitivity analyses (**Figure S4**). Cluster composition was subsequently confirmed if isolates shared a common ancestor according to ST^Pas^-specific core genome phylogeny (**Figure 3C-D**). All non-index and corresponding index isolates clustered closely (**Figure 3**), and comparable results were obtained when clustering by either ST^Pas^-specific accessory gene content or adjusting for potential recombination events (**Figure S5).**

**Figure 3.**
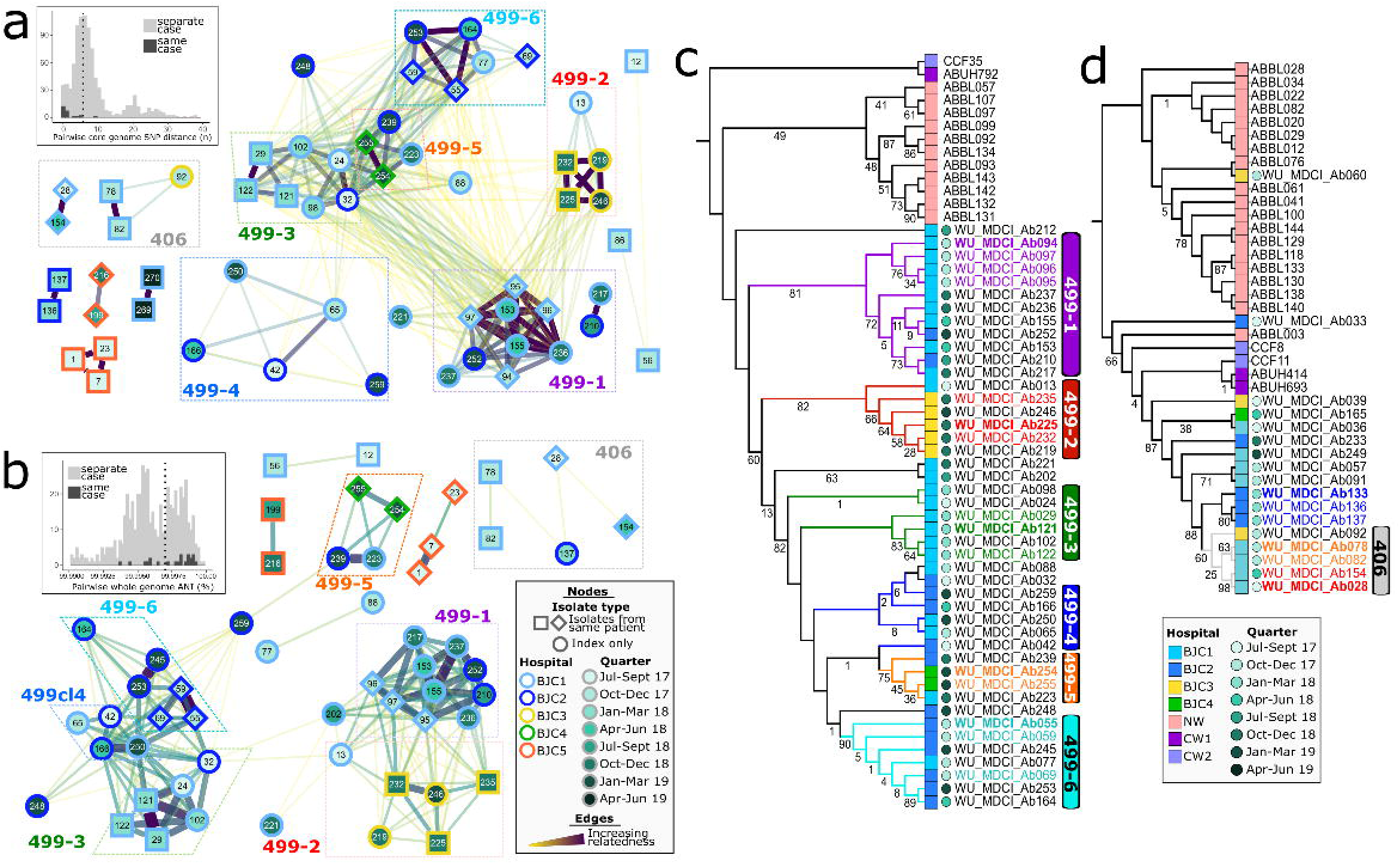
Clonal clusters were identified exclusively among ST499^Pas^ and ST406^Pas^ isolates. **(a,b)** Network analyses demonstrating relatedness of isolates according to core genome SNP distance (panel A) and whole genome ANI (panel B). Each node represents an isolate, with nodes color-coded according to isolation hospital (border) and calendar quarter (fill), per key in panel B. Edges represent interactions that meet cutoffs, with degree of relatedness represented by edge width and color; edge distances were manually adjusted for clarity. Insets demonstrate histograms of values from pairwise comparisons between isolates obtained from different patients (light gray) or isolates obtained from the same patient (dark gray). Only extreme values are displayed, for clarity. Dotted lines denote cutoff values for edges in respective networks. (**c)** Maximum likelihood tree derived using 1881 SNPs identified from alignment of 2·63 Mbp in 2702 core genes of ST499^Pas^ isolates. **(d)** Maximum likelihood trees derived using 16644 SNPs identified from alignment of 2·09 Mbp in 2110 core genes of ST406^Pas^ isolates. For clarity in panels C and D, only bootstrap values <90% are included. Each leaf is color-coded according to isolation hospital (square) and calendar quarter (circle), per key in panel D. Isolates obtained from the same patient share label color, with index isolates in bold. Isolates belonging to clonal clusters highlighted in panels A and B, are denoted by branch color and vertical labels in panel C and D.

We detected six ST499^Pas^ (**Figure 3C and 4**) and one ST406^Pas^ clonal clusters (**Figure 3D**), and suspected transmission sites were identified in four ST499^Pas^ clusters (**Figure 4C-F**). Cluster 499-1 was composed of four patients (A-D) with recent exposure to clinic 4 (CLIN4) and/or CLIN5; two overlapping residents of long-term care facility 6 (LTF6) patients colonized with the phylogenetically distinct isolates 210 and 217 (**Figure 4B,** patients D and E); and two patients with multiple incidences of ST499^Pas^/ast-5 isolates across many months (G, H), who had resided in LTF9 (**Figure 4C**). Patient 499-1F lacked an obvious link, except for recurrent admission to BJC1. Three of four cluster 499-2 patients repeatedly visited CLIN2 (**Figure 4D**), and all cluster 499-5 patients had resided in LTF10 during a 3-month span (**Figure 4E**). Lastly, 499-6 was the only cluster containing individuals (A-D) enrolled in the local Veterans Affairs (VA) medical system. Though VA records were not available, patients B-D were long-term residents of LTF26 (**Figure 4F**). Shared exposures were not identified for clusters 499-3, cluster 499-4 and 406, but no single hospital linked these cases, either. Moreover, patients 499-3B, 499-3D and 499-4D were not hospitalized in a study hospital prior to their index culture (**Figure 5**). In summary, various contemporary clonal networks were linked to cluster-specific, nonhospital settings that likely facilitated their propagation.

**Figure 4.**
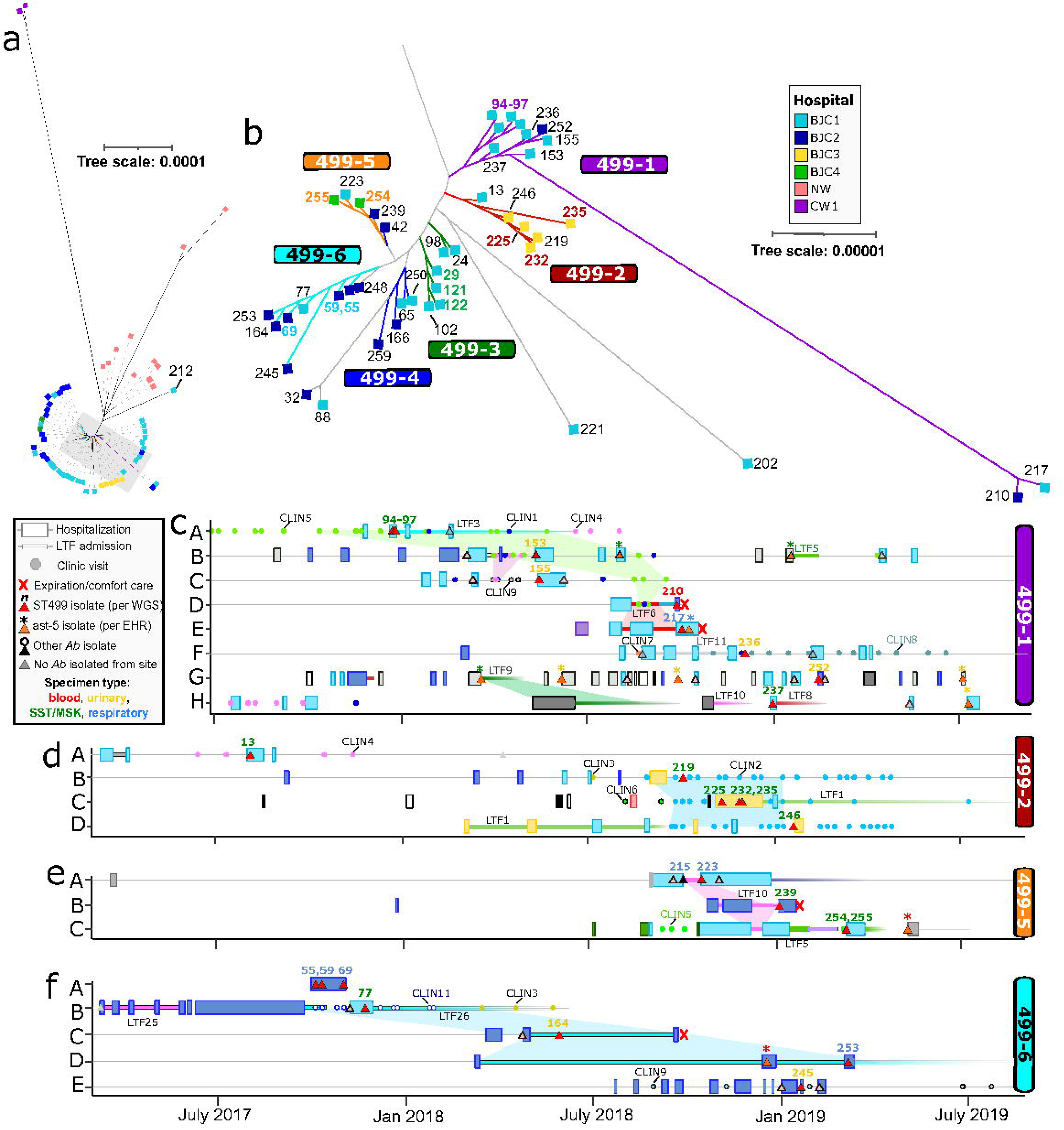
Epidemiologic links within ST499^Pas^ clonal clusters. (**a,b**) Unrooted version of ST499_p_ phylogenetic tree from Figure 3C. Panel B is ∼10x enlargement of region highlighted by gray rectangle in panel A. Clonal clusters are denoted by colored branches. Tree leaf symbols denote hospital of isolation (see panel B inset key). BJC isolates obtained from the same patient share label color. **(c-f)** Clinical timelines of patients (labeled A-H on Y-axis) in 499-1, 499-2, 499-5 and 499-6, respectively. As presented in the key adjacent to panel C, colored symbols on each patient timeline denote dates of hospitalizations (wide rectangles), long term care facilities admissions (LTF, narrow rectangles), outpatient clinic visits (CLIN, circles), and pertinent clinical cultures (triangles with labels colored according to culture specimen). Full color-coded key is in Figure 5. Shaded areas between timelines highlight shared exposures suspected of facilitating clonal propagation (see text). 28-month span represented on X-axis of panel F, is conserved in all panels. WGS, whole genome sequencing; EMR, electronic health record; *Ab*, *A. baumannii*.

**Figure 5.**
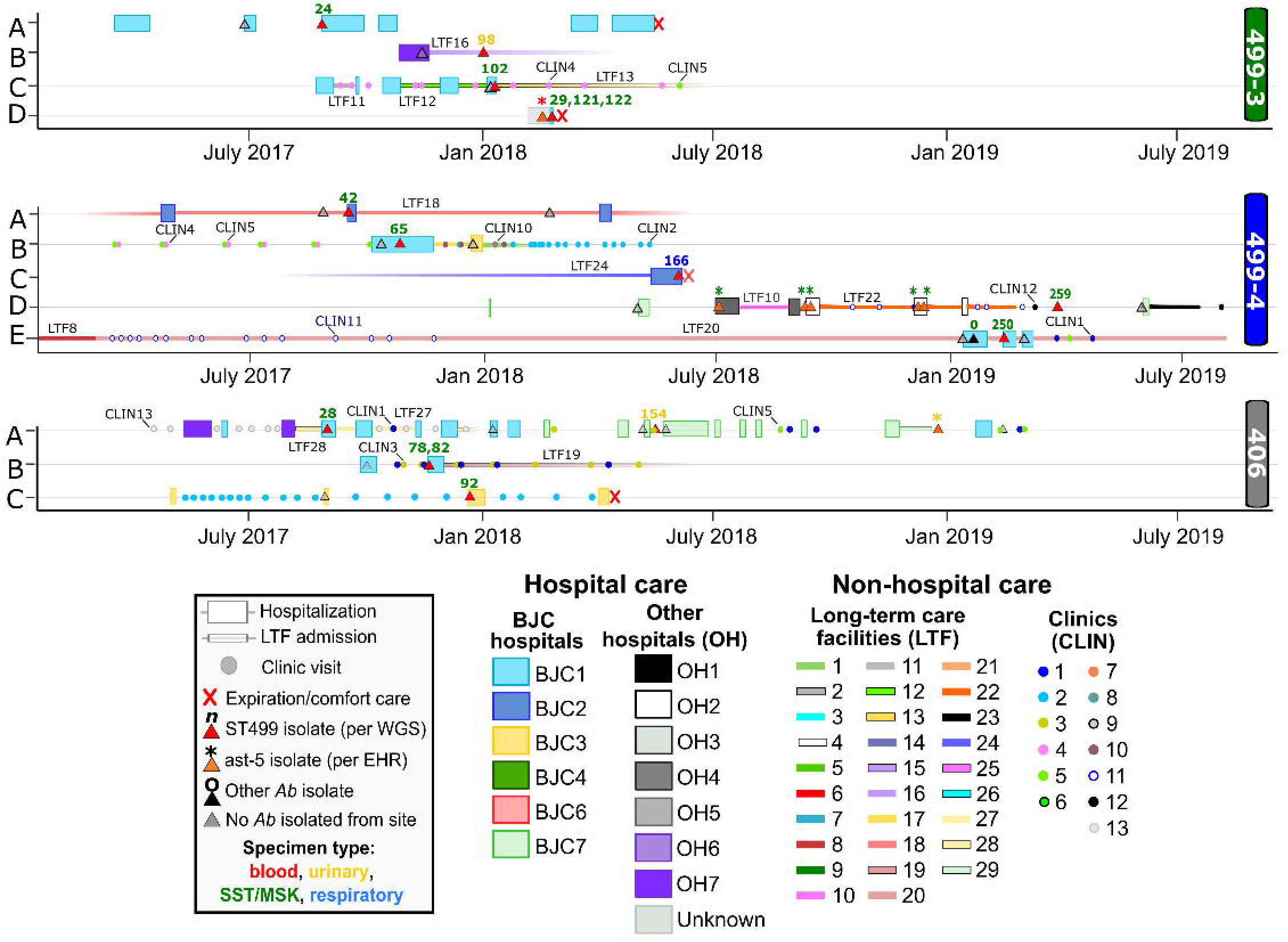
Patient timelines of clonal clusters lacking obvious epidemiologic links. Clinical timelines of patients (labeled A-E on Y-axis) in clonal clusters 449-3, cluster 499-4 and 406 (from top to bottom). Colored objects on each patient timeline (see key at bottom) denote dates of hospitalizations (wide rectangles), long-term care facilities admissions (LTF, narrow rectangles), outpatient clinic visits (CLIN, circles), and pertinent clinical cultures (triangles, labels colored according to culture specimen). 28-month span is conserved in all panels. Colored key also corresponds to patient timelines in Figure 4. WGS, whole genome sequencing; EMR, electronic medical record; *Ab*, *A. baumannii*.

## Discussion

We report sharp rise of ST499^Pas^ CR*Ab* cases in the St. Louis region, which remarkably coincided with the decline of CR*Ab* displaying differing antibiotypes. ST2^Pas^ and ST79^Pas^, two of the top three globally prevalent CR*Ab* lineages[17], overwhelmingly predominated *Ab* cases in recent molecular surveys at two other USA Midwest academic hospital systems (**Figure S3**)[24, 25], and evidence supports these lineages previously predominated in our region. The ast-1/-2 or ast-4 antibiotypes characteristic of BJC ST2^Pas^ or ST79^Pas^ isolates, respectfully (**Figure 2A**), were displayed by >85% of pre-2016 BJC CRAb isolates but only 17% of CR*Ab* in 2019. Furthermore, in a U.S. nationwide CR*Ab* survey conducted in 2008-2009, all isolates contributed by BJC were ST2^Pas^ or ST79^Pas^ (n=11 and 2, respectively)[19]. While these putative pre-2012 BJC ST2^Pas^ infections were overwhelmingly HA and arguably resulted from clonal, intrahospital outbreaks, recent ST2^Pas^ cases were seemingly result of sporadic, unrelated clones (**Figure S3**). Altogether, these findings imply the elimination of conditions that were once conducive to the propagation of major global lineages in the region.

Conversely, the coinciding emergence of BJC ST499^Pas^ reflects the lineage’s capacity to exploit a different epidemiological space. In contrast to CR*Ab* propagation centering on inpatient and critical care settings pre-2012, we appreciated no contribution from intrahospital transmission to the ongoing occurrence of ST499^Pas^ (**Figure 6**). Consistent with local CR*Ab* cases transitioning towards nHA SST/MSK and urinary cases[16], ST499^Pas^ propagation appeared to center around clinics serving patients with chronic wounds and LTFs (**Figure 4**). Though the latter are recognized hotspots for CR*Ab* transmission[6–8], wound clinics have been historically underappreciated. We observed multiple instances (e.g., patients 499-1B, 499-1G, 499-1H, 499-4D, etc.) in which ST499^Pas^ was isolated over the span of up to 12 months from a single patient, establishing that individuals residing in the community are either persistently colonized or repeatedly exposed to long-lived CR*Ab* pools outside of hospitals. This merits further investigation, as these asymptomatic individuals could serve as incidental vectors communicating outbreaks between different locations and exposing individuals at risk for severe disease[26].

**Figure 6.**
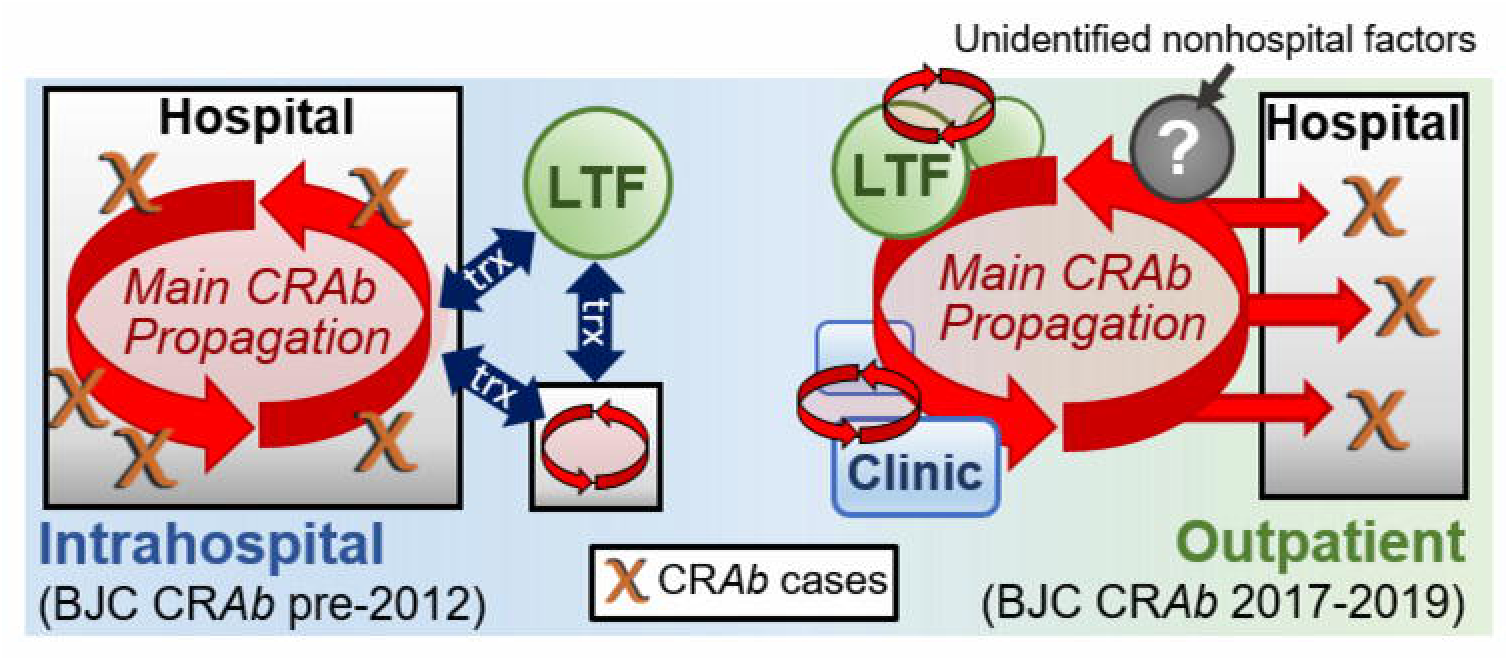
The spectrum of CR*Ab* propagation habits ranging from stable intrahospital pools (left) to centering around outpatient environments (right). LTF, long-term care facilities; trx, patient transfers.

Pasteur scheme ST499^Pas^ (not to be confused with the unrelated Oxford scheme ST499^Oxf^ [27]) was first described in the US in a 2010 blood isolate [28], although the single locus variant ST123^Pas^ was detected once among Las Vegas 2008-2009 isolates [19]. Remarkably, though the 2008-2009 U.S. nationwide survey identified no ST499 isolates [19], a subsequent analysis of CR*Ab* in four U.S. hospital centers (performed concurrently and independently from out study) revealed ST499^Pas^ as the second most prevalent lineage, second only to ST2^Pas^ [29]. ST499^Pas^ is not part of any major global clone and, apart from a three 2014 isolates in Tanzania and a one 2000 isolate in Australia, has been exclusively identified in North America[17, 30, 31]. Though capable of acquiring genes encoding broader resistance, as evidenced by isolate 212 (**Figure 2**), ST499^Pas^ displayed the most narrow CR*Ab* antibiotype, effectively dismissing antibiotics as the main driver of its emergence. Importantly, the fact that no ST499^Pas^ intrahospital outbreak has been reported to our knowledge, does not preclude its capacity to establish sustained intrahospital transmission and disease (akin to preceding lineages), if given the opportunity.

Though our study is limited to describing CR*Ab* in a single metropolitan area, BJC is representative of modern healthcare systems[24] where increasing reliance on outpatient services for managing chronic illnesses may facilitate outpatient CR*Ab* propagation. Surveillance studies remain the gold standard for tracking MDRO transmission trends, but non-targeted surveillance of outpatient environments can be resource intensive and complicated by local practice variations. Furthermore, the utility of surveillance screening of *Ab* has been brought into question[32], in part due to an incomplete understanding of *Ab* carriage and natural reservoirs. As shown here, combining genomic analysis of clinical isolates with a regionally integrated EMR system represents an alternative that can be especially valuable for guiding larger subsequent surveillance efforts.

Importantly, the resolution afforded by employing WGS and multilevel criteria for clonal clusters was key to disentangling CR*Ab* networks in a patient population with overlapping exposures. For example, patient 499-1H admission to LTF10 (**Figure 4A**) coincided with the LTF10 admission of cluster 499-5 patients (**Figure 4E**), but unambiguous assignment of isolate 237 to cluster 499-1, instead, implicates LTF9. Though our convenience cohort was representative of the overall BJC isolate population, being unable to assign shared exposures to some ST499^Pas^ clusters may have resulted from failure to capture keystone cases, including possible cases of intrahospital transmission. Also, future investigations should include other healthcare exposures associated with chronic medical conditions, including home health services, emergency room visits, non-wound/surgical clinics, and other outpatient services (e.g., radiology, dialysis, infusion centers, etc.). Lastly, the phylogenetic diversity of BJC Cs*Ab* isolates (**Figure 2A**) implies exposure to a deep *Ab* gene pool existing outside of healthcare settings and highlights that *Ab* is an environmental species that survives in various human-independent niches that remain to be identified[2]. The outpatient arena represents the next front in the fight against CR*Ab* disease, and elucidating how CR*Ab* persists in nonhospital settings will be a cornerstone for future combat strategies.

## Data Availability

All data produced in the present study are present in the manuscript, available online or upon reasonable request to the authors.  Sequence read files and constructed genomes for sequenced isolates are available on NCBI under BioProject PRJNA739144, and BioSample accession numbers SAMN19774044-4250. The datasets used and analyzed during the current study are available from JJC on reasonable request. Access to retrospective clinical case datasets including patient demographics and specific dates is restricted due to patient privacy protections. 

## Footnotes

### Prior presentations

This research was presented in part as an oral abstract (#154) at ID Week in October, 2020 and as an oral abstract at the AcinetoVibes Online Conference in June, 2022. **For reprints** please contact Gautam Dantas, 4515 McKinley Ave, Rm 5314, Box 8510, St Louis, MO 63110. [p] (314) 362-7238 [f] (314) 362-2156 [e].

### Authors’ change in affiliation

JJC currently is an Assistant Professor in the Division of Infectious Diseases, Department of Medicine, University of Alabama at Birmingham Heersink School of Medicine, Birmingham, Alabama, USA. Correspondence for JJC should be addressed to Bevill Biomedical Research Building, Room 534, 845 19th St S, Birmingham, AL 35205.

### Authors’ contributions

JJC conceived and designed the study. MCSA performed health record review. RFP was involved with design of comparative genomic analysis. CAB and MAW collected samples. JJC performed specimen handling, genome sequencing, data management and statistical analysis. JJC prepared figures and tables, and wrote the first draft of the manuscript. CAB and GD analyzed and discussed data and critically revised the manuscript.

## Acknowledgements

The authors thank the Edison Family Center for Genome Sciences & Systems Biology at WUSM staff, Eric Martin, Brian Koebbe, Jessica Hoisington-López, and MariaLynn Crosby for technical support in high-throughput sequencing and computing. The authors thank the members of the Dantas and Burnham lab for helpful comments and critiques. The authors thank Dorothy Sinclair and Cherie Hill for their essential and expert contributions in medical record data retrieval for this study.

## Ethics approval and consent to participate

This study was approved by the Institutional Review Board of Washington University in St. Louis (IRB# 201707046 and 201707049). A waiver of informed consent was granted as all specimens and clinical records were obtained during routine clinical care and many patients were already deceased or would otherwise have been unable to be contacted.

## Availability of data and materials

Sequence read files and constructed genomes for sequenced BJC isolates are available on NCBI under BioProject PRJNA739144, and BioSample accession numbers SAMN19774044-4250. The datasets used and analyzed during the current study are available from JJC on reasonable request. Access to retrospective clinical case datasets including patient demographics and specific dates is restricted due to patient privacy protections.

## Supplementary Tables

**Table S1.** List of clinical and epidemiological “elements” identified in electronic health record review. ^a^, shapes and abbreviations used in clinical timelines in Figures 4 and S9.

|  | Element type | Timeline key<sup>a</sup> | Definition | Collected information |
| --- | --- | --- | --- | --- |
| 1 | **Postive cultures** |  | **Cultures containing *Ab* or *Acbc* isolates of interest** | **Culture collection date** |
|  | ST406/ST409 isolate | ### [red triangle] | Cultures containing isolate obtained for WGS and typed by MLST | Anatomical and/or tissue source of culture |
|  | ast-3/ast-5 isolate | * [orange triangle] | Cultures containing isolate NOT obtained for WGS but identified by EHR | Anatomical and/or tissue source of culture |
| 2 | **Negative cultures** |  | **Cultures from same anatomical and/or tissue source as "positive cultures", but NOT containing *Ab* or *Acbc* isolates of interest** | **Culture collection date** |
|  | Other *Ab* isolate | O [black triangle] | Cultures containing *Ab* or *Acbc* isolates, but NOT of interests, according to MLST or EHR | Anatomical and/or tissue source of culture |
|  | No *Ab* isolate | [gray triangle] | Cultures containing non-*Acinetobacter* isolates or which grew no organisms | Anatomical and/or tissue source of culture |
| 3 | **Hospitalizations** | [colored box] | **Admission to hospital for >24 hours** | **Admission, hospital-to-hospital transfer (if pertinent) and discharge dates** |
|  | BJC | BJC1-7 | Admission to one of fifteen BJC hospitals in our region | Emergency room (ER) records and discharge summary, for origination and disposition information |
|  | Other hospital | OH1-7 | Admission to other hospitals, as noted in EHR | ER records and discharge summary, for origination and disposition information |
|  | Unknown hospital | "unknown" | Admission to/transfer from hospital, whose identity was not clearly stated in EHR |  |
|  | * NOTE: Emergency room and urgent care clinic encounters WITHOUT admission, were not included in final analysis |  |  |  |
| 4 | **Long-term care facility encounters** | LTF1-29 [colored bar] | **Admission to and/or residency in long-term care, long-term acute care, skilled nursing home, or assisted living facilities** | **Admission and discharge dates** |
|  | *NOTE: Unless clearly delineated in EHR, duration of LTF encounter was presumed according to preceding discharge records, subsequent admission records (i.e., whether patient was admitted from LTF or home), and outpatient records. |  |  |  |
| 5 | **Infectious disease clinic encounters** | CLIN1 [blue circle] | **In-person appointments at main BJC outpatient infectious disease clinic** | **Appointment date (excluded telemedicine and "no-show" appointments)** |
| 6 | **Wound care/surgical clinic encounters** | CLIN2-13 [colored circle] | **In-person appointments to any wound care specialty or general surgical outpatient clinic, as noted in EHR** | **Appointment date (excluded telemedicine and "no-show" appointments)** |
| 7 | **Expiration/comfort care** | X | **Person expired or decision to transition to comfort care** | **Date of expiration note, or date of progress note/discharge summary detailing transition to comfort care measures** |
| 8 | **Suspected transmission sites** | [shaded areas between patients] | either (1) an encounter for a patient colonized with a CR*Ab* isolate preceded the encounter of another patient who subsequently had a positive culture; or (2) encounters preceded positive culture for at least two patients in a cl. |  |

**Table S2.** Antibiotic resistance rates among CRAb isolates, BJC 2007-2019. p-values are comparing non-susceptibility rates of isolates obtained from 2007-12 versus 2013-19, by Fisher’s Exact Test. s, susceptible; ns, non-susceptible; NA, susceptibility data not available; %ns, percent non-susceptible

|  | Ciprofloxacin |  |  |  | Gentamicin |  |  |  | Trimethoprim/Sulfamethaxazole |  |  |  | Tetracycline and/or Doxycycline |  |  |  | Ampicillin/Sulbactam |  |  |  |
| --- | --- | --- | --- | --- | --- | --- | --- | --- | --- | --- | --- | --- | --- | --- | --- | --- | --- | --- | --- | --- |
|  | s | ns | NA | %ns | s | ns | NA | %ns | s | ns | NA | %ns | s | ns | NA | %ns | s | ns | NA | %ns |
| 2007 | 0 | 60 | 11 | 100.0 | 19 | 52 | 0 | 73.2 | 4 | 66 | 1 | 94.3 | 2 | 26 | 43 | 92.9 | 7 | 21 | 43 | 75.0 |
| 2008 | 0 | 102 | 25 | 100.0 | 54 | 73 | 0 | 57.5 | 5 | 122 | 0 | 96.1 | 27 | 99 | 1 | 78.6 | 25 | 101 | 1 | 80.2 |
| 2009 | 1 | 120 | 21 | 99.2 | 82 | 60 | 0 | 42.3 | 6 | 136 | 0 | 95.8 | 67 | 68 | 7 | 50.4 | 26 | 109 | 7 | 80.7 |
| 2010 | 0 | 85 | 24 | 100.0 | 70 | 39 | 0 | 35.8 | 3 | 106 | 0 | 97.2 | 55 | 53 | 1 | 49.1 | 73 | 28 | 8 | 27.7 |
| 2011 | 0 | 111 | 1 | 100.0 | 53 | 58 | 1 | 52.3 | 3 | 108 | 0 | 97.3 | 61 | 51 | 0 | 45.5 | 51 | 22 | 39 | 30.1 |
| 2012 | 0 | 44 | 0 | 100.0 | 10 | 34 | 0 | 77.3 | 4 | 40 | 0 | 90.9 | 20 | 24 | 0 | 54.5 | 29 | 14 | 1 | 32.6 |
| 2013 | 0 | 53 | 0 | 100.0 | 22 | 31 | 0 | 58.5 | 12 | 40 | 1 | 76.9 | 23 | 29 | 1 | 55.8 | 13 | 40 | 0 | 75.5 |
| 2014 | 2 | 43 | 0 | 95.6 | 27 | 18 | 0 | 40.0 | 6 | 39 | 0 | 86.7 | 22 | 23 | 0 | 51.1 | 17 | 28 | 0 | 62.2 |
| 2015 | 1 | 57 | 0 | 98.3 | 37 | 19 | 2 | 33.9 | 10 | 48 | 0 | 82.8 | 19 | 38 | 1 | 66.7 | 30 | 24 | 4 | 44.4 |
| 2016 | 0 | 44 | 0 | 100.0 | 22 | 22 | 0 | 50.0 | 9 | 34 | 1 | 79.1 | 14 | 8 | 22 | 36.4 | 23 | 15 | 6 | 39.5 |
| 2017 | 0 | 30 | 0 | 100.0 | 17 | 13 | 0 | 43.3 | 4 | 26 | 0 | 86.7 | 27 | 3 | 0 | 10.0 | 19 | 11 | 0 | 36.7 |
| 2018 | 0 | 40 | 0 | 100.0 | 27 | 13 | 0 | 32.5 | 13 | 27 | 0 | 67.5 | 36 | 4 | 0 | 10.0 | 29 | 11 | 0 | 27.5 |
| 2019 | 2 | 51 | 0 | 96.2 | 47 | 6 | 0 | 11.3 | 27 | 26 | 0 | 49.1 | 49 | 4 | 0 | 7.5 | 37 | 16 | 0 | 30.2 |
| **TOTAL 2007-12** | **1** | **522** | **82** | **99.8** | **288** | **316** | **1** | **52.3** | **25** | **578** | **1** | **95.9** | **232** | **321** | **52** | **58.0** | **211** | **295** | **99** | **58.3** |
| **TOTAL 2013-19** | **5** | **318** | **0** | **98.5** | **199** | **122** | **2** | **38.0** | **81** | **240** | **2** | **74.8** | **190** | **109** | **24** | **36.5** | **168** | **145** | **10** | **46.3** |
| **p =** | **0.0327** |  |  |  | **<0.0001** |  |  |  | **<0.0001** |  |  |  | **<0.0001** |  |  |  | **0.0009** |  |  |  |

**Table S3.**
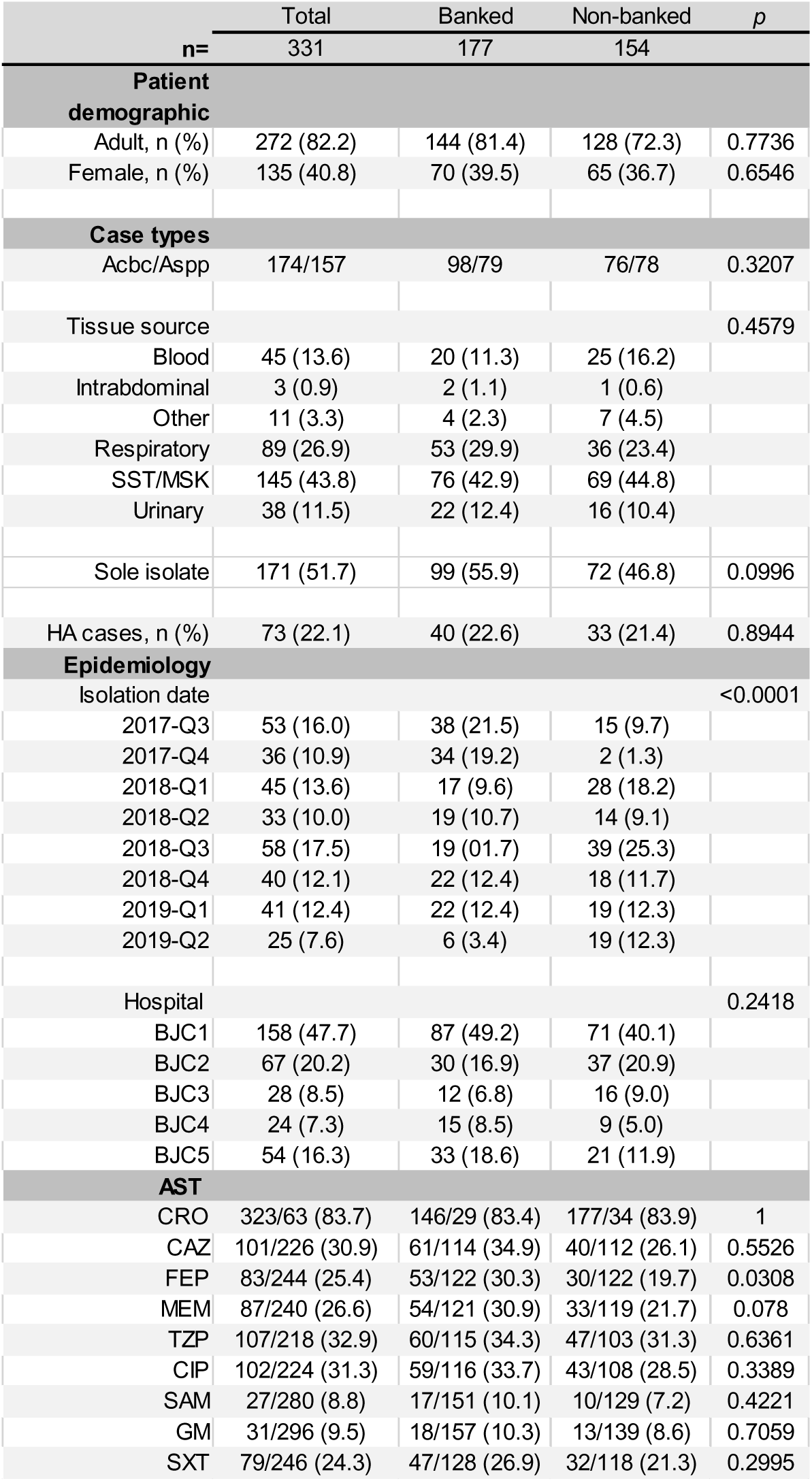
Clinical characteristics *Acinetobacter* cases with banked and non-banked isolates, BJC 7/2017-5/2019. ^a^, p-value comparing genotyped to not genotyped cases, according to Fisher’s exact test. The proportion of isolates begonging to >2 categories, was compared for “tissue source”, “isolation date” and “hospital”. ^b^, Cases lacking AST results in the EMR were excluded from analysis.

| n= | Total<br>331 | Banked<br>177 | Non-banked<br>154 | p |
| --- | --- | --- | --- | --- |
| **Patient demographic** |  |  |  |  |
| Adult, n (%) | 272 (82.2) | 144 (81.4) | 128 (72.3) | 0.7736 |
| Female, n (%) | 135 (40.8) | 70 (39.5) | 65 (36.7) | 0.6546 |
| **Case types** |  |  |  |  |
| Acbc/Aspp | 174/157 | 98/79 | 76/78 | 0.3207 |
| Tissue source |  |  |  | 0.4579 |
| Blood | 45 (13.6) | 20 (11.3) | 25 (16.2) |  |
| Intrabdominal | 3 (0.9) | 2 (1.1) | 1 (0.6) |  |
| Other | 11 (3.3) | 4 (2.3) | 7 (4.5) |  |
| Respiratory | 89 (26.9) | 53 (29.9) | 36 (23.4) |  |
| SST/MSK | 145 (43.8) | 76 (42.9) | 69 (44.8) |  |
| Urinary | 38 (11.5) | 22 (12.4) | 16 (10.4) |  |
| Sole isolate | 171 (51.7) | 99 (55.9) | 72 (46.8) | 0.0996 |
| HA cases, n (%) | 73 (22.1) | 40 (22.6) | 33 (21.4) | 0.8944 |
| **Epidemiology** |  |  |  |  |
| Isolation date |  |  |  | <0.0001 |
| 2017-Q3 | 53 (16.0) | 38 (21.5) | 15 (9.7) |  |
| 2017-Q4 | 36 (10.9) | 34 (19.2) | 2 (1.3) |  |
| 2018-Q1 | 45 (13.6) | 17 (9.6) | 28 (18.2) |  |
| 2018-Q2 | 33 (10.0) | 19 (10.7) | 14 (9.1) |  |
| 2018-Q3 | 58 (17.5) | 19 (10.7) | 39 (25.3) |  |
| 2018-Q4 | 40 (12.1) | 22 (12.4) | 18 (11.7) |  |
| 2019-Q1 | 41 (12.4) | 22 (12.4) | 19 (12.3) |  |
| 2019-Q2 | 25 (7.6) | 6 (3.4) | 19 (12.3) |  |
| Hospital |  |  |  | 0.2418 |
| BJC1 | 158 (47.7) | 87 (49.2) | 71 (40.1) |  |
| BJC2 | 67 (20.2) | 30 (16.9) | 37 (20.9) |  |
| BJC3 | 28 (8.5) | 12 (6.8) | 16 (9.0) |  |
| BJC4 | 24 (7.3) | 15 (8.5) | 9 (5.0) |  |
| BJC5 | 54 (16.3) | 33 (18.6) | 21 (11.9) |  |
| **AST** |  |  |  |  |
| CRO | 323/63 (83.7) | 146/29 (83.4) | 177/34 (83.9) | 1 |
| CAZ | 101/226 (30.9) | 61/114 (34.9) | 40/112 (26.1) | 0.5526 |
| FEP | 83/244 (25.4) | 53/122 (30.3) | 30/122 (19.7) | 0.0308 |
| MEM | 87/240 (26.6) | 54/121 (30.9) | 33/119 (21.7) | 0.078 |
| TZP | 107/218 (32.9) | 60/115 (34.3) | 47/103 (31.3) | 0.6361 |
| CIP | 102/224 (31.3) | 59/116 (33.7) | 43/108 (28.5) | 0.3389 |
| SAM | 27/280 (8.8) | 17/151 (10.1) | 10/129 (7.2) | 0.4221 |
| GM | 31/296 (9.5) | 18/157 (10.3) | 13/139 (8.6) | 0.7059 |
| SXT | 79/246 (24.3) | 47/128 (26.9) | 32/118 (21.3) | 0.2995 |

**Table S4.**
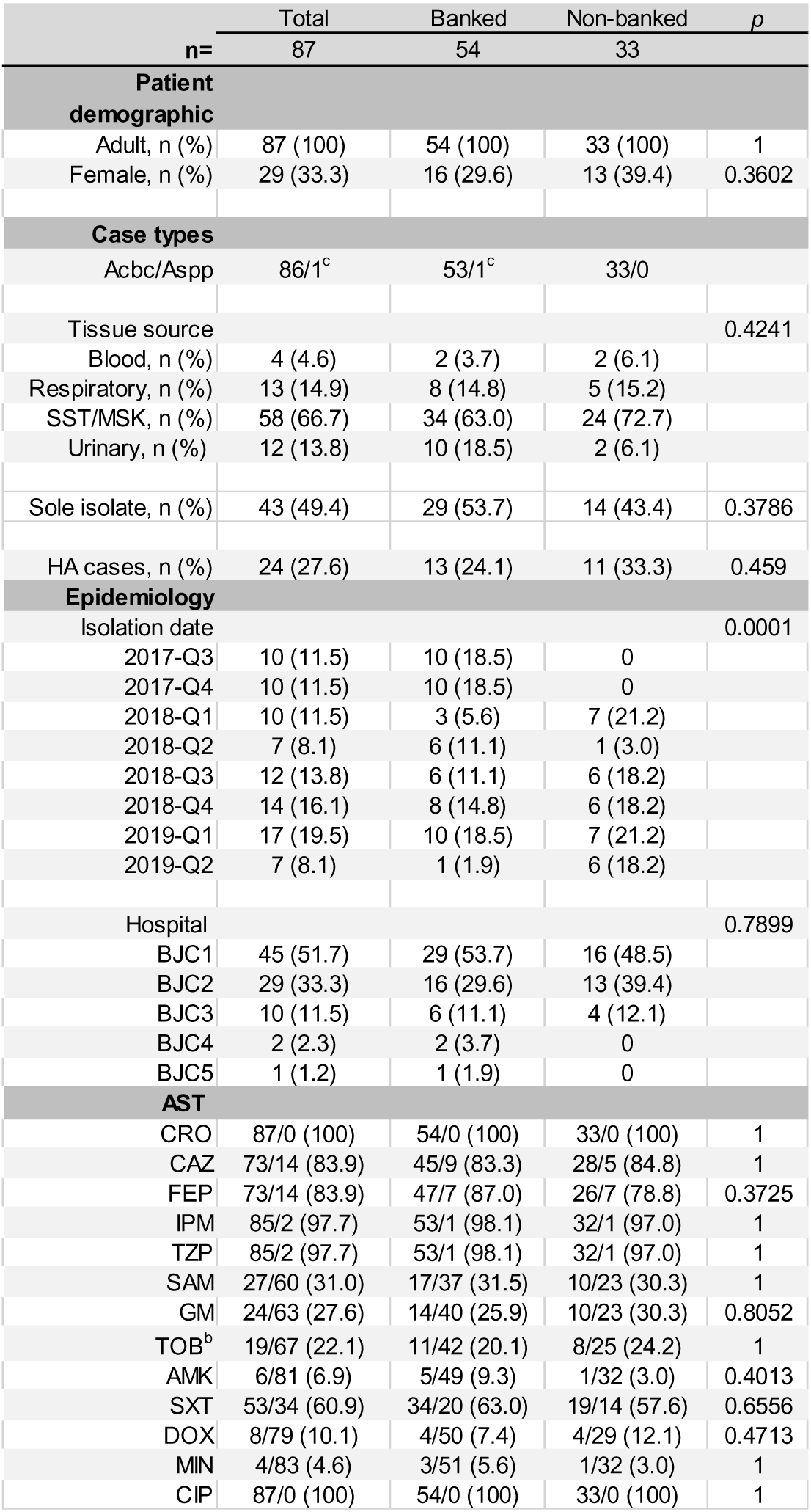
Clinical characteristics CR*Ab* cases with banked and non-banked isolates, BJC 7/2017-5/2019. ^a^, p-value comparing genotyped to not genotyped cases, according to Fisher’s exact test. The proportion of isolates begonging to >2 categories, was compared for “tissue source”, “isolation date” and “hospital”. ^b^, One genotyped isolate was missing TOB susceptibility data in the EMR. ^c^, isolate initially reported as Aspp was actually *A. baumannii* by sequencing analysis.

**Table S5.**
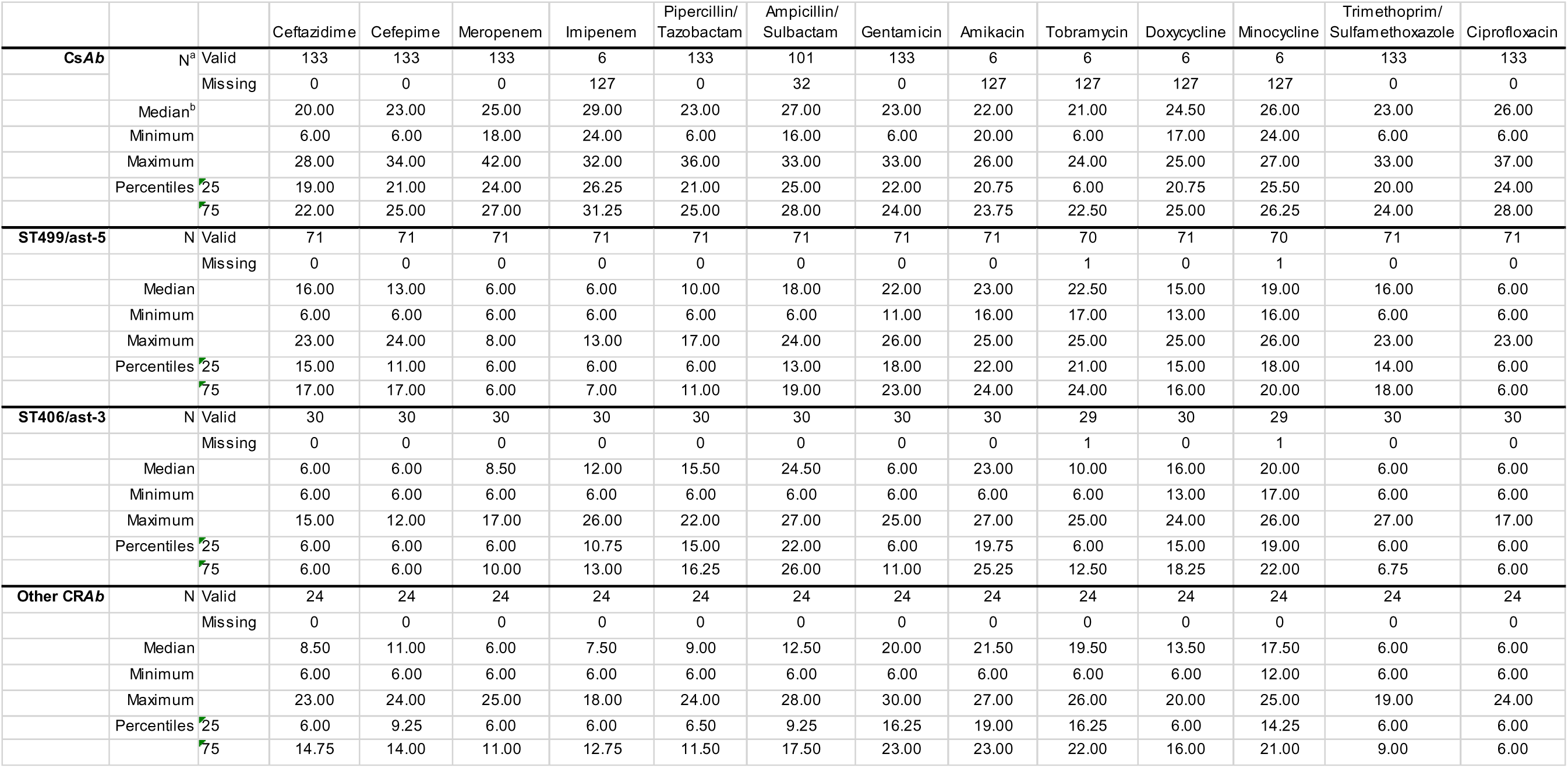
Summary of AST data for each group of *A. baumannii* isolates from Figures 2b-d and S2b. ^a^, number of isolates with valid or missing susceptibility testing data for each antibiotic; ^b^, All median, minimum, maximum and percentile values are zone of clearance sizes, mm

|  |  |  | Ceftazidime | Cefepime | Meropenem | Imipenem | Piperacillin/<br>Tazobactam | Ampicillin/<br>Sulbactam | Gentamicin | Amikacin | Tobramycin | Doxycycline | Minocycline | Trimethoprim/<br>Sulfamethoxazole | Ciprofloxacin |
| --- | --- | --- | --- | --- | --- | --- | --- | --- | --- | --- | --- | --- | --- | --- | --- |
| **CsAb** | N<sup>a</sup> | Valid | 133 | 133 | 133 | 6 | 133 | 101 | 133 | 6 | 6 | 6 | 6 | 133 | 133 |
|  |  | Missing | 0 | 0 | 0 | 127 | 0 | 32 | 0 | 127 | 127 | 127 | 127 | 0 | 0 |
|  | Median<sup>b</sup> |  | 20.00 | 23.00 | 25.00 | 29.00 | 23.00 | 27.00 | 23.00 | 22.00 | 21.00 | 24.50 | 26.00 | 23.00 | 26.00 |
|  | Minimum |  | 6.00 | 6.00 | 18.00 | 24.00 | 6.00 | 16.00 | 6.00 | 20.00 | 6.00 | 17.00 | 24.00 | 6.00 | 6.00 |
|  | Maximum |  | 28.00 | 34.00 | 42.00 | 32.00 | 36.00 | 33.00 | 33.00 | 26.00 | 24.00 | 25.00 | 27.00 | 33.00 | 37.00 |
|  | Percentiles | 25 | 19.00 | 21.00 | 24.00 | 26.25 | 21.00 | 25.00 | 22.00 | 20.75 | 6.00 | 20.75 | 25.50 | 20.00 | 24.00 |
|  |  | 75 | 22.00 | 25.00 | 27.00 | 31.25 | 25.00 | 28.00 | 24.00 | 23.75 | 22.50 | 25.00 | 26.25 | 24.00 | 28.00 |
| **ST499/ast-5** | N | Valid | 71 | 71 | 71 | 71 | 71 | 71 | 71 | 71 | 70 | 71 | 70 | 71 | 71 |
|  |  | Missing | 0 | 0 | 0 | 0 | 0 | 0 | 0 | 0 | 1 | 0 | 1 | 0 | 0 |
|  | Median |  | 16.00 | 13.00 | 6.00 | 6.00 | 10.00 | 18.00 | 22.00 | 23.00 | 22.50 | 15.00 | 19.00 | 16.00 | 6.00 |
|  | Minimum |  | 6.00 | 6.00 | 6.00 | 6.00 | 6.00 | 6.00 | 11.00 | 16.00 | 17.00 | 13.00 | 16.00 | 6.00 | 6.00 |
|  | Maximum |  | 23.00 | 24.00 | 8.00 | 13.00 | 17.00 | 24.00 | 26.00 | 25.00 | 25.00 | 25.00 | 26.00 | 23.00 | 23.00 |
|  | Percentiles | 25 | 15.00 | 11.00 | 6.00 | 6.00 | 6.00 | 13.00 | 18.00 | 22.00 | 21.00 | 15.00 | 18.00 | 14.00 | 6.00 |
|  |  | 75 | 17.00 | 17.00 | 6.00 | 7.00 | 11.00 | 19.00 | 23.00 | 24.00 | 24.00 | 16.00 | 20.00 | 18.00 | 6.00 |
| **ST406/ast-3** | N | Valid | 30 | 30 | 30 | 30 | 30 | 30 | 30 | 30 | 29 | 30 | 29 | 30 | 30 |
|  |  | Missing | 0 | 0 | 0 | 0 | 0 | 0 | 0 | 0 | 1 | 0 | 1 | 0 | 0 |
|  | Median |  | 6.00 | 6.00 | 8.50 | 12.00 | 15.50 | 24.50 | 6.00 | 23.00 | 10.00 | 16.00 | 20.00 | 6.00 | 6.00 |
|  | Minimum |  | 6.00 | 6.00 | 6.00 | 6.00 | 6.00 | 6.00 | 6.00 | 6.00 | 6.00 | 13.00 | 17.00 | 6.00 | 6.00 |
|  | Maximum |  | 15.00 | 12.00 | 17.00 | 26.00 | 22.00 | 27.00 | 25.00 | 27.00 | 25.00 | 24.00 | 26.00 | 27.00 | 17.00 |
|  | Percentiles | 25 | 6.00 | 6.00 | 6.00 | 10.75 | 15.00 | 22.00 | 6.00 | 19.75 | 6.00 | 15.00 | 19.00 | 6.00 | 6.00 |
|  |  | 75 | 6.00 | 6.00 | 10.00 | 13.00 | 16.25 | 26.00 | 11.00 | 25.25 | 12.50 | 18.25 | 22.00 | 6.75 | 6.00 |
| **Other CRAb** | N | Valid | 24 | 24 | 24 | 24 | 24 | 24 | 24 | 24 | 24 | 24 | 24 | 24 | 24 |
|  |  | Missing | 0 | 0 | 0 | 0 | 0 | 0 | 0 | 0 | 0 | 0 | 0 | 0 | 0 |
|  | Median |  | 8.50 | 11.00 | 6.00 | 7.50 | 9.00 | 12.50 | 20.00 | 21.50 | 19.50 | 13.50 | 17.50 | 6.00 | 6.00 |
|  | Minimum |  | 6.00 | 6.00 | 6.00 | 6.00 | 6.00 | 6.00 | 6.00 | 6.00 | 6.00 | 6.00 | 12.00 | 6.00 | 6.00 |
|  | Maximum |  | 23.00 | 24.00 | 25.00 | 18.00 | 24.00 | 28.00 | 30.00 | 27.00 | 26.00 | 20.00 | 25.00 | 19.00 | 24.00 |
|  | Percentiles | 25 | 6.00 | 9.25 | 6.00 | 6.00 | 6.50 | 9.25 | 16.25 | 19.00 | 16.25 | 6.00 | 14.25 | 6.00 | 6.00 |
|  |  | 75 | 14.75 | 14.00 | 11.00 | 12.75 | 11.50 | 17.50 | 23.00 | 23.00 | 22.00 | 16.00 | 21.00 | 9.00 | 6.00 |

## Supplementary Figure Legends

**Figure S1.**
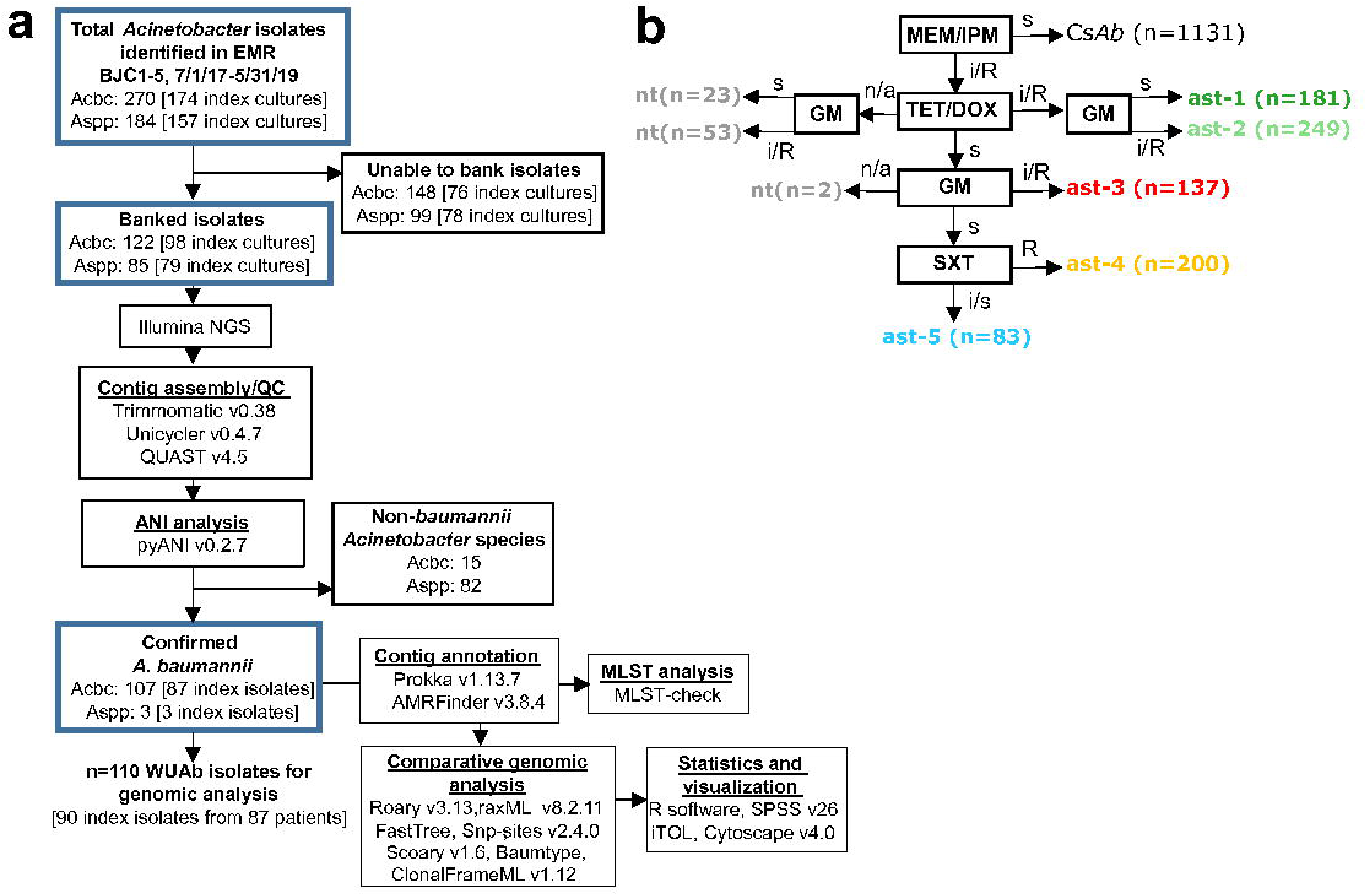
Analysis workflows. **(a)** Overview of banking and processing of *Acinetobacter* clinical isolates collected between July 2017 and May 2019. All isolates were reported as *Acinetobacter calcoaceticus-baumannii complex* (Acbc) or *Acinetobacter species* (Aspp) according to MALDI-TOF performed in the BJC clinical microbiology laboratory. (**b)** Algorithm for assigning antibiotype according to antimicrobial susceptibility testing (AST). All BJC CR*Ab* index cultures isolates (n=928) were typed ast-1 through ast-5 according to whether they were reported as resistant (R), intermediate (i), or susceptible (s) to listed antibiotics. If an isolate’s AST result was not available (n/a) for any antibiotic in the algorithm, it was classified as non-typeable (nt). The number of index cultures in each category is noted in parentheses. GM, gentamicin; MEM/IPM, meropenem and/or imipenem; SXT, trimethoprim/sulfamethaxazole; TET/DOX, tetracycline and/or doxycycline.

**Figure S2.**
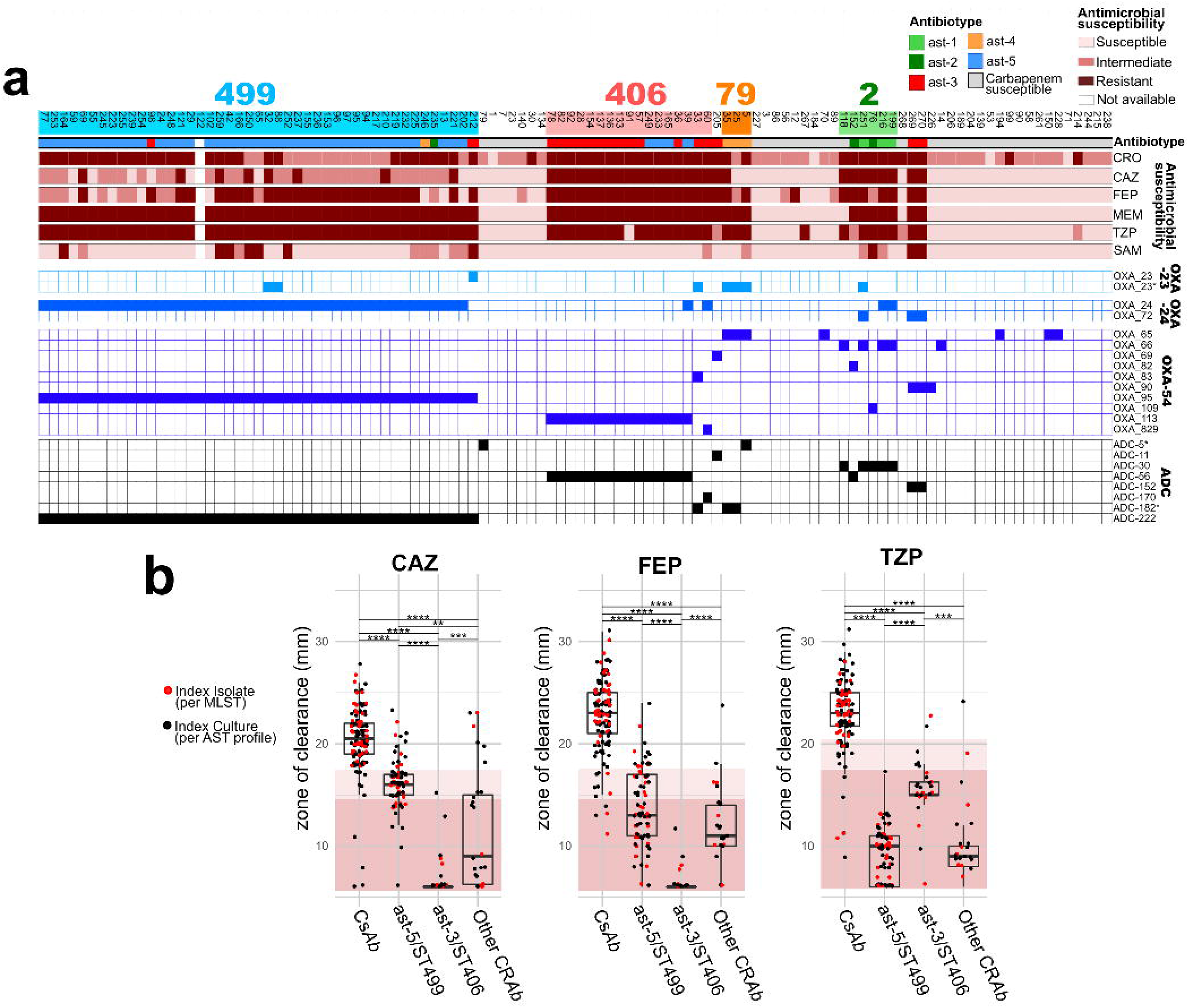
BJC CRAb isolate beta-lactamase gene content and beta-lactam AST. **(a)** ST^Pas^ containing CRAb isolates are highlighted and labeled. First colored row denotes antibiotype, and subsequent rows denote AST results to beta-lactam antibiotics routinely tested with all *Acinetobacter* isolates in the BJC CML. Bottom boxes denote presence/absence of beta-lactamase genes, listed on right and grouped according to homologous lactamase types (bold). For clarity, only genes identified in CRAb isolates are included. Isolate metadata are listed in **Data S2**, and all identified resistance genes are listed in **Data S4**. **(b)** AST results according to zone of clearance, of genotyped *Ab* index isolates (red dots) and non-genotyped Acbc index cultures (black dots) identified in BJC hospitals between January 2017 and December 2019. Isolates are grouped according to antibiotype. Backgrounds highlight ranges for “resistant” (dark) and “intermediate” (light) susceptibility, per CLSI guidelines. Box-plot center lines denote medians, while box limits denote upper and lower quartile values (listed in Table S5). Whiskers denote 1.5x interquartile range. Medians compared according to Mann-Whitney test with Bonferroni adjustment for multiple comparisons. *, p<0·05; **, p<0·005; ***, p<0·0005; ****, p<0·00005.

**Figure S3.**
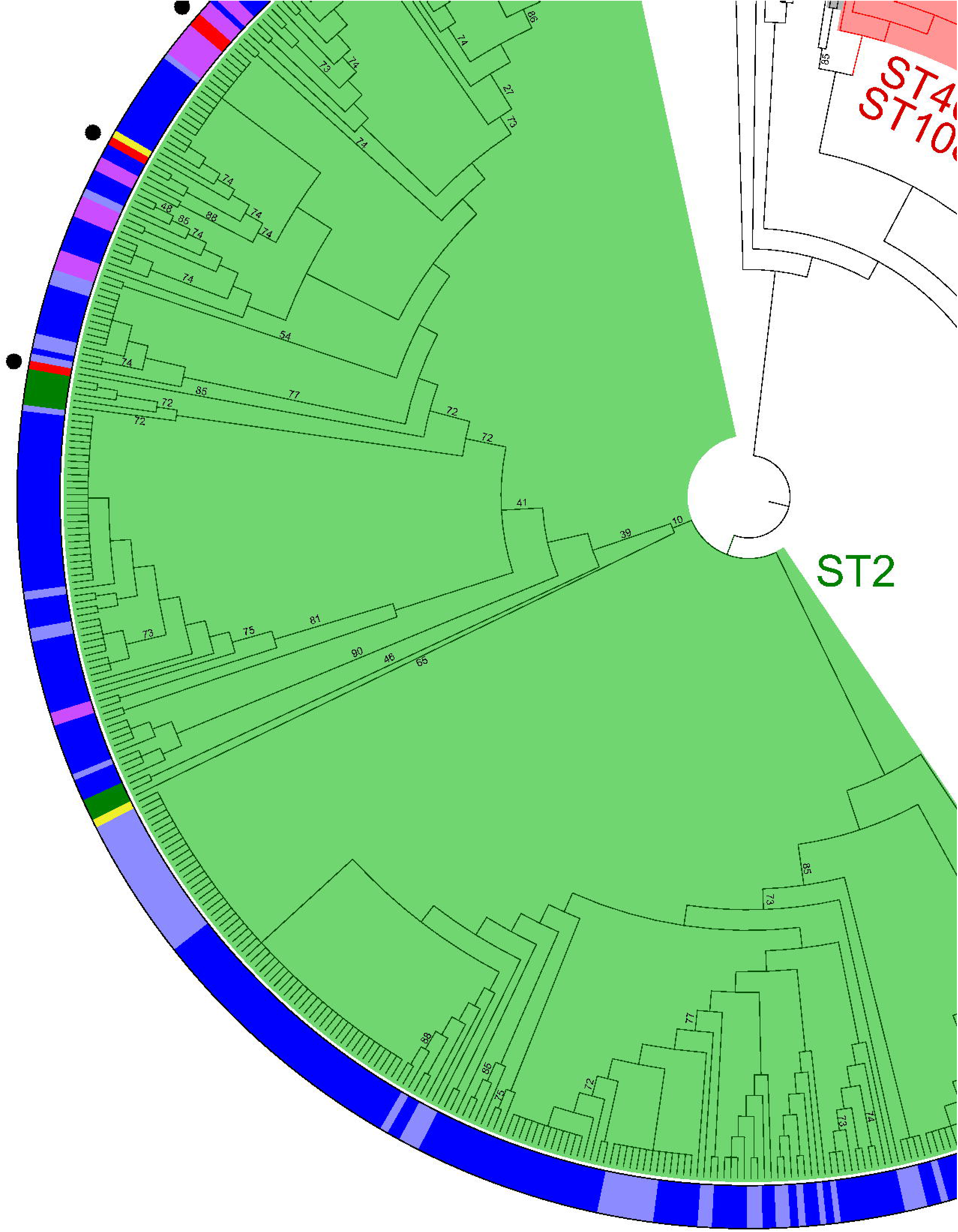
Global core genome phylogeny of *A. baumannii* isolates. Maximum likelihood phylogenetic tree comparing 1647 core genes shared by select *A. baumannii* genomes available on NCBI and BJC isolates. The outer ring depicts the medical center in which the isolate was identified. Clades corresponding to the predominant ST^Pas^ are color coded and labeled. ST406^Pas^ and ST1088^Pas^ are single locus variants. Black dots denote separate phylogenetic groups of BJC ST2^Pas^ and ST406^Pas^ isolates. For clarity, only bootstrap values ≤90% are included on corresponding branches.

**Figure S4.**
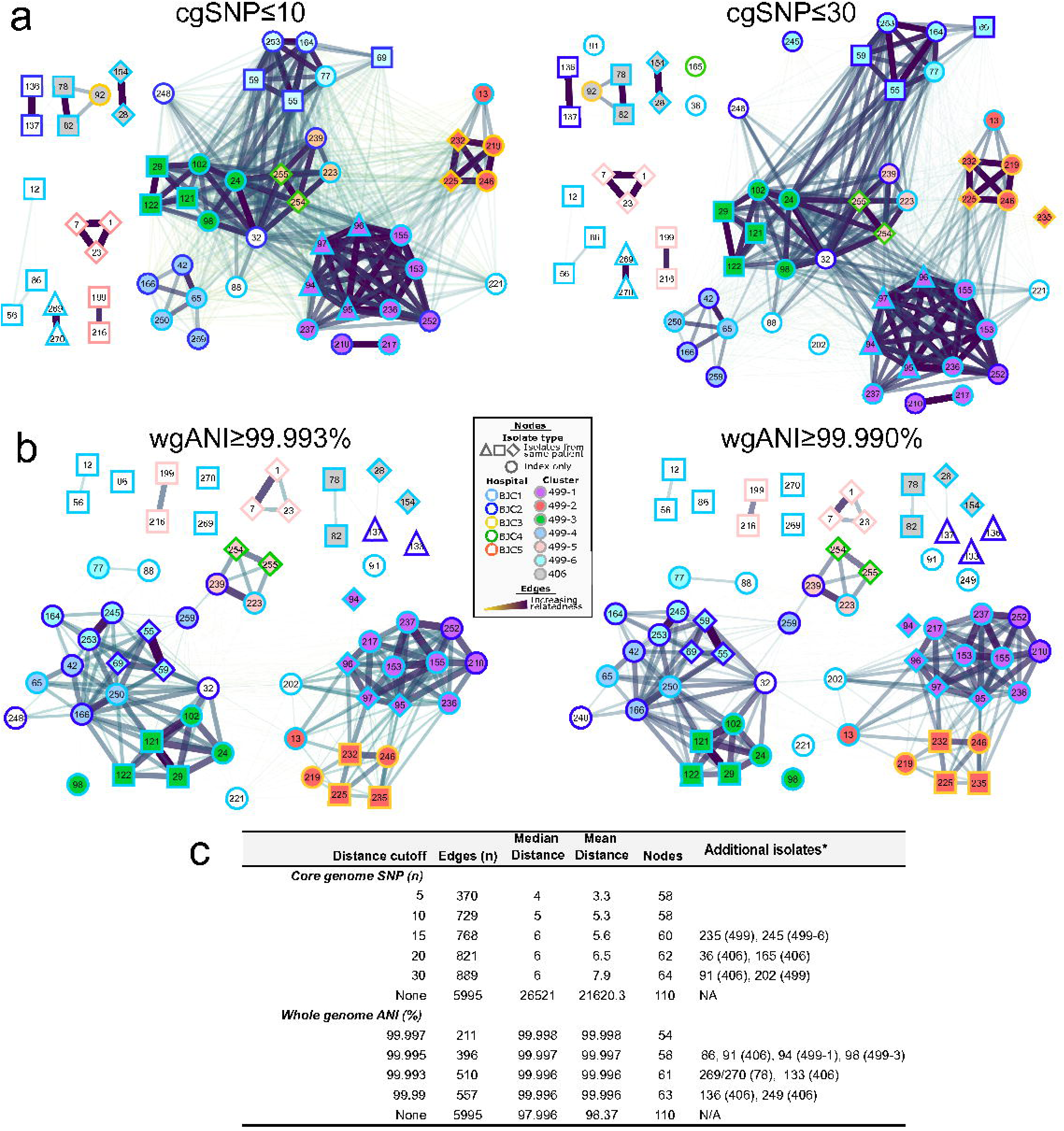
Sensitivity analyses of criteria for assigning presumptive clonal cluster. **(a,b)** Networks demonstrating relatedness of isolates according to different cutoffs (listed above networks) for core genome SNP distance (cgSNP, panel a) and whole genome ANI (wgANI, panel b). Each node represents an isolate, with nodes color-coded according to isolation hospital (border) and clonal cluster according to **Figure 3** (fill), per key in panel B. Edges represent interactions that meet cutoffs, with degree of relatedness represented by edge width and color. Node positions and edge distances were manually adjusted for better comparison with F**igure 3** and clarity. **(c)** Table listing network changes as result from adjusting the cutoff values. “Additional isolates*” column lists the additional isolates (and corresponding sequence type/clonal cluster in parentheses) that are eligible for inclusion compared to one step stricter cutoffs.

**Figure S5.**
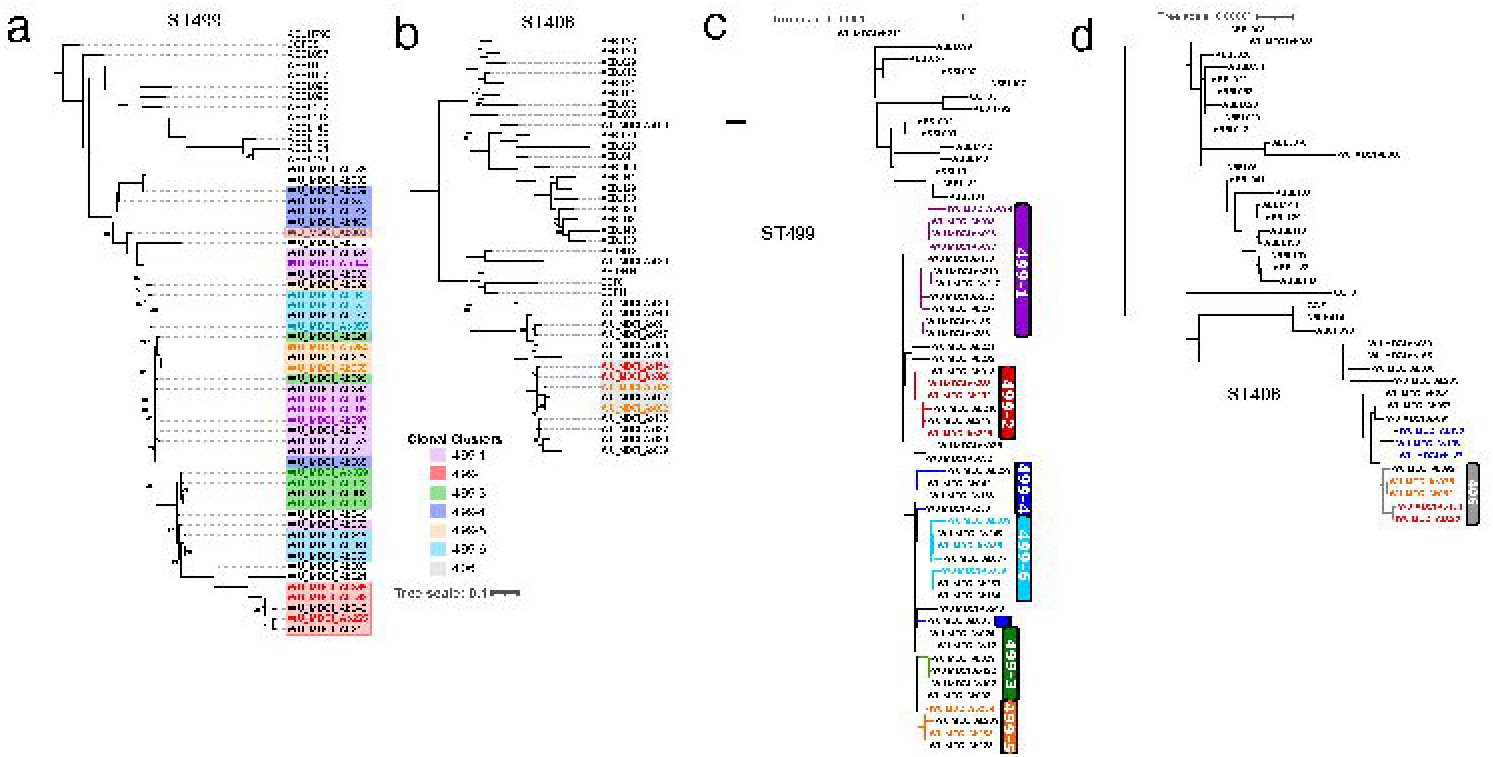
Confirmation of ST499^Pas^ and ST406^Pas^ phylogeny by alternative methodology. (**a,b)** Maximum likelihood phylogenetic trees according to the presence-absence of accessory genes in ST499^Pas^ (panel a) and ST406^Pas^ (panel b) isolates. For clarity, only bootstrap values ≤90% are included on corresponding branches. Isolates obtained from the same patient share label color, with index isolates in bold. Tree scale in key applies to both trees. **(c,d)** Maximum likelihood phylogenetic trees according to the core genome alignment of ST499^Pas^ (panel c) and ST406^Pas^ (panel d) isolates, corrected for computationally predicted recombination events using ClonalFrameML software. Isolates obtained from the same patient share label color, with index isolates in bold. Isolates belonging to clonal clusters as identified in **Figure 3**, are highlighted by color or denoted by branch color and vertical labels. Isolate WU_MDCI_Ab065 is assigned to cluster 499-4.

## Supplementary Data

**Data S1 (DataS1_BJCAcbcEpi.xls)**

Three tables containing raw data of epidemiological analysis of *Acbc* cases, BJC 2007-2019.

**Data S2 (DataS2_BJC_IsolateList.xls)**

Table listing name, sequence information and clinical metadata of BJC isolates included in genomic analysis.

**Data S3 (DataS3_NCBIisolates.xls)**

Table containing list of NCBI genomes and corresponding isolates, included in this study.

**Data S4 (DataS4_BJC_IsolateARGanalysis.xls)**

Results of genomic analysis for antimicrobial resistance genes and other genomic element, BJC isolates.

