## Supplemental Text for "Outpatient clonal propagation propelled rapid regional establishment of an emergent carbapenem-resistant *Acinetobacter baumannii* lineage ST499^Pas^"

**Supplementary methods description**

**Methods used for whole genome sequencing and sequencing analysis.**

Isolates were propagated on sheep’s blood agar, and *Acinetobacter* isolate genomic DNA was extracted with the Gentra Puregene Yeast/Bact. Kit (Qiagen, Germantown, MD, USA) according to manufacturer’s instructions. A total of 5 ng/uL of DNA was used as input for Illumina sequencing libraries with the Nextera kit (Illumina, San Diego, CA, USA). Pooled libraries were sequenced on a NextSeq HighOutput platform (Illumina) to obtain 2 x 150 bp reads. Following demultiplexing by barcode, reads had adapters removed with Trimmomatic v0.38 [1]. Reads were then assembled into draft genomes using de-novo assembler Unicycler v0.4.7 [2]. Scaffolds .fasta files were used for downstream analysis. Assembly statistics was quantified using QUAST v4.5 [3].

**Genomic analysis.**

Species identity of each genome was confirmed using the ANIm method from pyANI v0.2.7[4]. ANIm ≥96% compared to the *A. baumannii* reference genome (strain 19606, GCF_002811175.1) was used as the species cutoff. In silico screen for antibiotic resistance genes and multilocus sequence typing (MLST) was performed with AMRFinder v3.8.4[5] and MLST-check[6], respectively. Prokka v1.13.7[7] was run on scaffold files to identify open reading frames >500 bp in length.

**Phylogenetic analysis.**

We performed three levels of phylogenetic analysis on *Ab* genomes: level 1) genomes of all BJC isolates and those obtained from NCBI (**Data S1**); level 2) genomes of all BJC isolates, only; and level 3) genomes of all BJC and NCBI isolates belonging to a single ST (i.e., ST499^Pas^ and ST406^Pas^). Using, gff files produced by Prokka (above) and Roary v3.13[8] with the script “roary -e -n -p [number of genomes] -i 95 *.gff”, we identified the core and accessory genomes (determined according to 95% nucleotide identity) for each level of analysis. The resulting “core_gene_alignment.fasta” file was used to generate a maximum likelihood tree (MLt) with FastTree v2.1.9[9] (for level 1) or raxML v8.2.11[10] (for levels 2 and 3). The “accessory_binary_genes.fa.newick” file produced by Roary was used to construct MLt based on the presence and absence of accessory genes. All MLts were visualized along with respective metadata, using iTOL[11]. Using the “core_gene_alignment.fasta” from level 2 phylogenetic analysis, Snp-sites v2.4.0[12] was used to remove indels and create multiFASTA alignment containing the core genome single nucleotide polymorphism (SNP) sites for each core genome.

**Clonality analysis.**

Clonal clusters were defined among BJC isolates using empirically-derived criteria and a multi-tiered approach. First, we calculated pairwise core genome SNP distance (from snp-sites multiFASTA alignment, above) and total genome average nucleotide identity (ANI, using pyANI results, above). The relatedness of index and non-index isolates obtained from the same patient served as reference to empirically determine a cutoff to define a clonal relationship between isolates obtained from different individuals. Networks of interactions between isolates meeting the cutoff were visualized using Cytoscape v4.0[13]. Isolates were assigned to “presumptive” clonal clusters if they demonstrated high genomic sequence similarity (defined as an interaction greater than median ANI or less than median SNP distance, within respective networks) and/or shared interactions with ≤5 other isolates in visualized networks. Lastly, MLt were constructed by aligning core genomes of isolates within a single ST (level 3 analysis, above). “Presumptive” clonal cluster isolates that shared a common ancestor with a bootstrap value ≥75% were assigned to a “confirmed” clonal cluster. Confirmed clonal clusters (cl) were labeled with their respective ST and an incremental number (i.e., “499-1”), and patients in each cl were labeled according to when a pertinent isolate was identified in their clinical timeline, with patient A having the earliest collection date.

**Supplementary Text References**
